# Prevalence of Antibodies to SARS-CoV-2 following natural infection and vaccination in Irish Hospital Healthcare Workers; changing epidemiology as the pandemic progresses

**DOI:** 10.1101/2021.11.04.21265921

**Authors:** Niamh Allen, Melissa Brady, Una Ni Riain, Niall Conlon, Lisa Domegan, Antonio Isidro Carrion Martin, Cathal Walsh, Lorraine Doherty, Eibhlin Higgins, Colm Kerr, PRECISE Study Steering Group, Colm Bergin, Catherine Fleming

## Abstract

**Background:** In October 2020 SARS-CoV-2 seroprevalence among hospital healthcare workers (HCW) of two Irish hospitals was 15% and 4.1% respectively. We compare seroprevalence in the same HCW population six months later, assess changes in risk factors for seropositivity with progression of the pandemic and serological response to vaccination.

**Methods:** All staff of both hospitals (N=9038) were invited to participate in an online questionnaire and SARS-CoV-2 antibody testing in April 2021. We measured anti-nucleocapsid and anti-spike antibodies. Frequencies and percentages for positive SARS-CoV-2 antibodies were calculated and adjusted relative risks for participant characteristics were calculated using multivariable regression analysis.

**Results:** 5085 HCW participated. Seroprevalence increased to 21% and 13% respectively; 26% of infections were previously undiagnosed. Black ethnicity (aRR 1.7, 95% CI 1.3-2.2, p<.001), lower level of education (aRR 1.4 for secondary level education, 95% CI 1.1-1.8, p=0.002), living with other HCW (aRR 1.2, 95% CI 1.0-1.4, p=0.007) were significantly associated with seropositivity. Having direct patient contact also carried a significant risk (being a healthcare assistant (aRR 1.8, 95% CI 1.3-2.3, p<.001), being a nurse (aRR 1.4, 95% CI 1.1-1.5, p=0.022), daily contact with COVID-19 patients (aRR 1.4, 95% CI 1.1-1.7, p=0.002), daily contact with patients without suspected or confirmed COVID-19 (aRR 1.3, 95% CI 1.1- 1.5, p=0.013) Breakthrough infection occurred in 23/4111(0.6%) of fully vaccinated participants; all had anti-S antibodies.

**Conclusion:** The increase in seroprevalence reflects the magnitude of the third wave of the pandemic in Ireland. Genomic sequencing is needed to apportion risk to the workplace versus the household/community. Concerted efforts are needed to mitigate risk factors due to ethnicity and lower level of education, even at this stage of the pandemic. The undiagnosed and breakthrough infections call for ongoing infection prevention and control measures and testing of HCW in the setting of close contact. Vaccinated HCW with confirmed infection should be actively assessed, including SARS-CoV-2 whole genome sequencing (WGS), serology testing and assessment of host determinants, to advance understanding of the reasons for breakthrough infection.

## Introduction

### SARS-CoV-2 infection in hospital healthcare workers

Healthcare workers, and those they live with, are at increased risk of contracting SARS-CoV-2 viral infection (1) (2) (3). Detectable antibody to SARS-CoV-2 is an excellent indicator of previous SARS-CoV-2 infection (4). A high proportion of the SARS-CoV-2 infections notified worldwide have been in hospital healthcare workers (HCW) and antibody seroprevalence has been shown to be higher in HCW than in the general population (5) (6) (7). Understanding the transmission and potential immunity dynamics of SARS-CoV-2 in hospitals is important in mitigating transmission at hospital level and adds valuable information to the growing evidence on the transmission patterns of COVID-19 among HCW.

### Antibody response following infection and vaccination

Natural infection has been shown to produce humoral and cellular immunity and whilst this may decline over time, durable memory responses are seen (8) (9). Vaccines have been shown to be protective both against infection and against symptomatic disease (10) (11) (12) (13). Vaccine-induced immunity produces a more robust response in the adaptive immune system therefore vaccination is likely to produce a sustained immune response and protection, including against currently known variants of concern (VoC) (14) (15). Robust B and T cell responses to vaccination have been shown for both mRNA vaccines and viral vector vaccines (16). Antibody response has been shown to correlate with protective immunity against infection (17).

The spike (S) and nucleocapsid (N) proteins are two of the main immunogens of the coronavirus proteins (18). Commercial SARS-CoV-2 antibody assays can detect antibodies to these structural proteins. Natural infection with SARS-CoV-2 elicits antibodies against the spike protein and the nucleocapsid protein (19). Currently available vaccines against the SARS-CoV-2 virus target the spike protein only (20) (21). The detection of anti-N antibodies allows vaccine-induced seroconversion to be distinguished from antibodies elicited by natural infection (22).

### Study sites

Hospital 1 is a tertiary referral hospital in the south inner city of Dublin, the capital city of Ireland (population 1.2 million) and has almost 4,700 employees and just over 1000 beds. It is one of the largest acute hospitals in Dublin city. Hospital 2 is a comparable tertiary referral hospital with almost 4400 employees and over 500 beds, located in Galway, in the West of Ireland (population 80,000). It is the main acute hospital serving the city of Galway. Both hospitals received patients with COVID-19 infection throughout the first wave of the pandemic in Ireland, and breakdown by ward and specialty is similar. The community incidence of COVID-19 in County Galway was significantly lower than in County Dublin during the first and second waves of the pandemic in Ireland (23). The first part of this study was conducted in October 2020, during the second wave of the pandemic in Ireland, and prior to the roll-out of COVID-19 vaccination. It showed an overall SARS-CoV-2 seroprevalence of 15% in Hospital 1 and 4.1% in Hospital 2 respectively. Almost 40% of infections had been previously undiagnosed (6) (24). The HCW seroprevalence was six times higher than community seroprevalence (5). During the third wave of the pandemic in Ireland (peak January 2021) the 14-day incidence for Galway approached that of Dublin (25). By the start of April 2021 the cumulative incidence of PCR-confirmed infections in HCW in Hospital 1 and 2 had risen to 18.5% and 9.2% respectively.

The purpose of this repeat cross-sectional study was to re-assess the prevalence of anti-SARS-CoV-2 antibodies in HCW in these two hospitals following the third, and largest, wave of the pandemic in Ireland, and assess changes in demographic, living arrangements and work-related risk factors. We also aimed to assess serological response to COVID-19 vaccination in the vaccinated sub-group, and to quantify breakthrough infections post vaccination.

## Materials and Methods

### Study Design and participants

This was a cross-sectional study of the seroprevalence of circulating antibodies to SARS-CoV-2, carried out from the 19^th^-28^th^ April 2021. All staff members of both hospitals (N=9038) were invited to participate in an online self-administered consent process and online questionnaire, followed by blood sampling for SARS-CoV-2 antibody testing in April 2021, in the same manner as October 2020 (6). Electronic consent and patient reported outcomes were captured using an eClinical platform Castor. (26). Information collected in the questionnaire included demographic information, contact details, place and type of work, level of contact with patients, previous COVID-19 symptoms and testing, history of close contact with a confirmed case of COVID-19, living arrangements and history of COVID-19 vaccination, including dates and type of vaccine. Blood samples were processed anonymously. Results were sent by text message to all participants on an opt-out basis. Results were discussed in person with any participant who requested this.

All vaccinated study participants received their COVID-19 vaccine as part of a two-dose regimen of the Comirnaty (Pfizer/BioNTech) vaccine, the Vaxzevria (formerly AstraZeneca) vaccine or the Moderna vaccine. A participant was considered partially vaccinated at ≥14 days after receipt of the first dose of vaccination, and fully vaccinated ≥14 days after receipt of the second dose of vaccination in line with Irish and international guidelines (27, 28).

### Laboratory Methods

All samples were tested using the Roche Elecsys anti-SARS-CoV-2 and the Roche Elecsys anti-SARS-CoV-2 S immunoassays detecting total antibodies (including IgG) to the nucleocapsid and spike proteins of the SARS-CoV-2 virus, respectively (29). Thresholds for positive results were as per manufacturers’ guidelines (29) (30). Participants with detectable anti-N antibodies were presumed to have had previous natural infection. Participants with detectable anti-S antibodies, and no reported history of COVID-19 vaccination were also presumed to have had natural infection. Participants with detectable anti-S antibodies and a history of COVID-19 vaccination were presumed to have these anti-S antibodies in response to vaccination.

### Statistical analysis

Frequencies and percentages were calculated for sociodemographic, epidemiological, and clinical characteristics. Participants were deemed seropositive (i.e. assumed to have had past infection with SARS-CoV-2) if they had detectable anti-N antibodies, or if they had detectable anti-S antibodies but had not been previously vaccinated. Characteristics of those who were seropositive were compared to those who were not seropositive, using the chi-square test. Univariable logistic regression was used to calculate relative risks along with their 95% confidence intervals to assess the association between SARS-CoV-2 seropositivity and characteristics of the study participants. Multivariable logistic regression analysis was conducted to control for negative and positive confounding and to calculate adjusted relative risks (aRR). No explicit finite population correction or reweighting was carried out. All analysis was conducted in Stata 15.1 (StataCorp LCC. 2019. College Station, TX 77845: USA).

### Ethical approval

Ethical approval was obtained from the National Research Ethics Committee (NREC) for COVID-19, Study Number 20-NREC. COV-101 (33).

### Funding

This work was supported financially by the Irish Health Service Executive COVID-19 budget.

## Results

### 1. SARS-CoV-2 seroprevalence (past infection)

#### Participation rates and demographics

In total 5,085 HCW participated (56% of invited staff). These 5,085 participants included 3,313 HCW who also participated in the first phase. On combined hospital data, 78% of participants were female. Median age was 40 years (IQR 30-49). By ethnicity 75% of participants were white Irish, 12% were Asian, and 2.3% were of African or any other black background. Age and sex of participants were similar in both hospitals while ethnicity differed slightly (Table 1a). Ninety-one percent of participants lived with other people and 31% lived with other HCW. The highest proportion (37%) of participants were nursing staff, 21% were allied healthcare staff, 14% were doctors, 13% were administration staff, 7.2% were general support staff and 5.7% health care assistants (HCA). Participation by staff grouping was similar in both hospitals and participants’ working roles broadly reflected the overall breakdown by role of the staff in both hospitals. (For details on participation by HCW role see Table A-D, Annex, main PRECISE 2 Study Report (31)).

**Table 1a.** Participant characteristics by hospital and total number of participants, April 2021

| Participant characteristics |  | Hospital 1 |  | Hospital 2 |  | P-value* | Total<br>(N=5,085) |  |
| --- | --- | --- | --- | --- | --- | --- | --- | --- |
|  |  | (N=2,945) |  | (N=2,140) |  |  |  |  |
|  |  | n | % | n | % |  | n | % |
| Age groups | 18-29 | 653 | 22 | 455 | 21 | 0.431 | 1,108 | 22 |
|  | 30-39 | 765 | 26 | 565 | 26 |  | 1,330 | 26 |
|  | 40-49 | 811 | 28 | 603 | 28 |  | 1,414 | 28 |
|  | 50-59 | 565 | 19 | 386 | 18 |  | 951 | 19 |
|  | ≥60 | 151 | 5.1 | 131 | 6.1 |  | 282 | 5.5 |
| Sex | Female | 2,278 | 77 | 1,681 | 79 | 0.309 | 3,959 | 78 |
|  | Male | 667 | 23 | 459 | 21 |  | 1,126 | 22 |
| Ethnicity | Irish | 2,091 | 71 | 1,707 | 80 | <0.001 | 3,798 | 75 |
|  | Any other white background | 257 | 8.7 | 219 | 10 |  | 476 | 9.4 |
|  | Asian background | 470 | 16 | 129 | 6.0 |  | 599 | 12 |
|  | African/other black background | 69 | 2.3 | 48 | 2.2 |  | 117 | 2.3 |
|  | Other | 58 | 2.0 | 37 | 1.7 |  | 95 | 1.9 |
| Country of birth | Ireland | 2,025 | 69 | 1605 | 75 | <0.001 | 3,630 | 71 |
|  | United Kingdom | 134 | 4.6 | 161 | 7.5 |  | 295 | 5.8 |
|  | India | 225 | 7.6 | 68 | 3.2 |  | 293 | 5.8 |
|  | Philippines | 198 | 6.7 | 16 | 0.7 |  | 214 | 4.2 |
|  | Poland | 26 | 0.9 | 59 | 2.8 |  | 85 | 1.7 |
|  | USA | 21 | 0.7 | 34 | 1.6 |  | 55 | 1.1 |
|  | Other | 316 | 11 | 197 | 9.2 |  | 513 | 10 |
| Education | Primary | 20 | 0.7 | 2 | 0.1 | <0.001 | 22 | 0.4 |
|  | Secondary | 409 | 14 | 200 | 9.3 |  | 609 | 12 |
|  | Third level | 1,280 | 43 | 964 | 45 |  | 2,244 | 44 |
|  | Post-graduate | 1,236 | 42 | 974 | 46 |  | 2,210 | 43 |
| Role | Admin | 403 | 14 | 273 | 13 | <0.001 | 676 | 13 |
|  | Medical/dental | 357 | 12 | 356 | 17 |  | 713 | 14 |
|  | Nursing/ midwifery | 1097 | 37 | 794 | 37 |  | 1,891 | 37 |
|  | Allied health | 612 | 21 | 432 | 20 |  | 1,044 | 21 |
|  | General support | 243 | 8.3 | 122 | 5.7 |  | 365 | 7.2 |
|  | Health care assistant | 179 | 6.1 | 112 | 5.2 |  | 291 | 5.7 |
|  | Other | 54 | 1.8 | 51 | 2.4 |  | 105 | 2.1 |
| Lives with | Alone | 270 | 9.2 | 194 | 9.1 | 0.603 | 464 | 9.1 |
|  | With others | 2,667 | 90. | 1,943 | 90.8 |  | 4,610 | 90.7 |
|  | Missing | 8 | 0.3 | 3 | 0.1 |  | 11 | 0.2 |
| Lives with HCW | Yes | 928 | 32 | 643 | 30 | 0.284 | 1,571 | 31 |
|  | No | 1,964 | 67 | 1,448 | 68 |  | 3412 | 67 |
|  | Missing | 53 | 1.8 | 49 | 2.3 |  | 102 | 2.0 |

\*Calculated using the chi-square test

#### Previous exposure, symptoms and testing

Hospital 1 staff had a higher percentage of previously confirmed SARS-CoV-2 infection; 18% of participants in Hospital 1 and 14% of participants in Hospital 2 reported that they had previously tested positive for COVID-19 infection by PCR. Table 1b shows the COVID-19 related characteristics of the participants by hospital.

**Table 1b.**
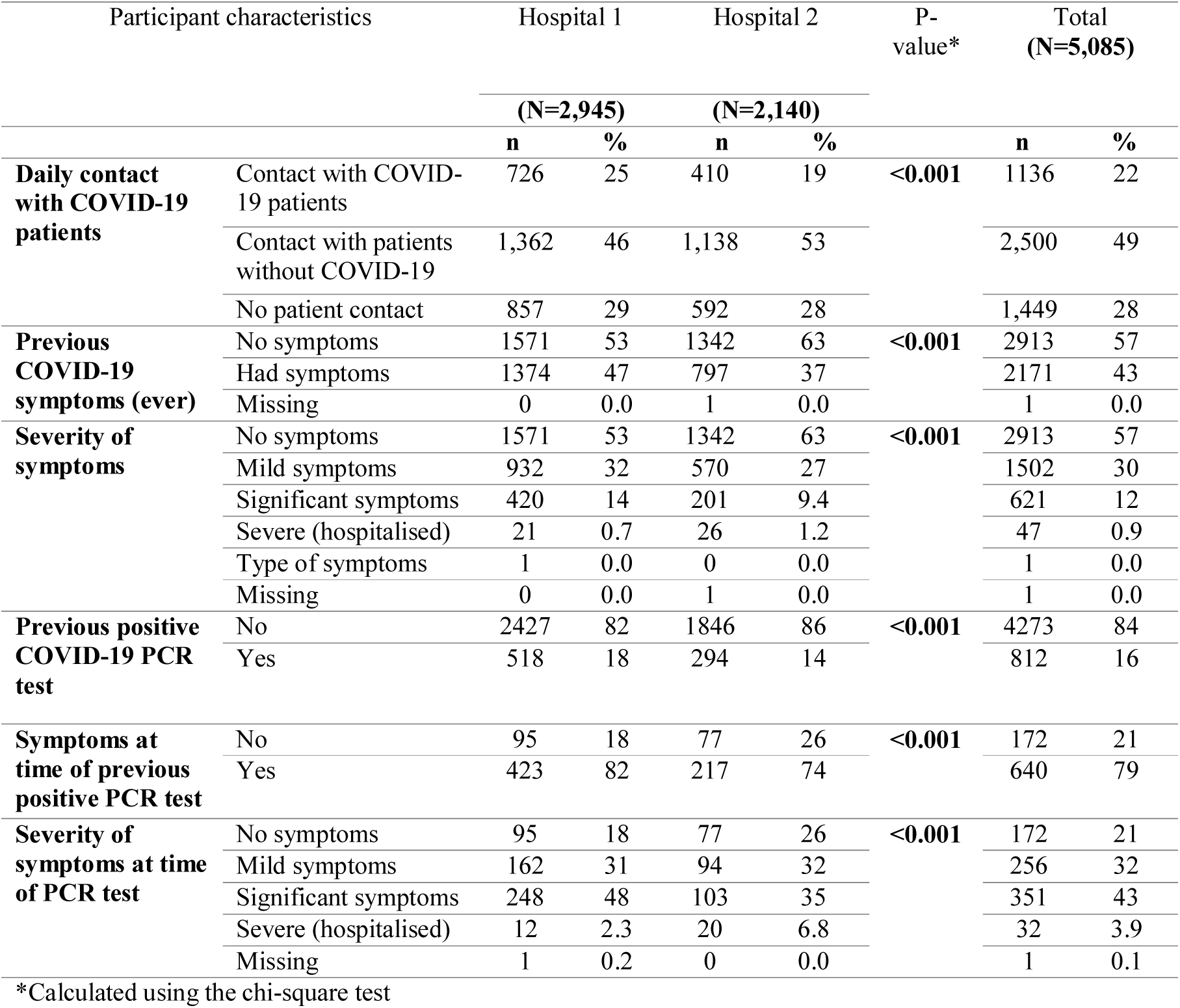
COVID-19 related characteristics by hospital and total number of participants, April 2021

#### Seroprevalence of antibodies to SARS-CoV-2

The overall seroprevalence of antibodies to SARS-CoV-2, indicative of past natural infection, was 21% in Hospital 1 and 13% in Hospital 2. Seroprevalence was higher in those with direct patient contact (especially those working with patients with COVID-19 infection).

Seroprevalence by degree of patient contact, sociodemographic characteristics and COVID-19 characteristics are shown in Table 2a and Table 2b. (Breakdown by hospital is shown in Tables 2c-f, Annex). By professional subgroup, the highest seroprevalence was seen amongst HCAs (32%), followed by general support staff (22%) and nurses/midwives (21%). (A detailed breakdown of general support staff by hospital is shown in Table 2g, Annex).

**Table 2a.** Prevalence of SARS-CoV-2 seropositivity by participant characteristics, both hospitals, April 2021

| Participant characteristics |  | Total | SARS-CoV-2 seropositive |  |  |
| --- | --- | --- | --- | --- | --- |
|  |  | N | n | % (95% CI) | p-value* |
| **Overall** |  | 5085 | 898 | 18 (17 - 19) |  |
| **Hospital** | Hospital 1 | 2,945 | 623 | 21 (20 - 23) | **<0.001** |
|  | Hospital 2 | 2,140 | 275 | 13 (11 - 14) |  |
| **Age groups (years)** | 18-29 | 1108 | 249 | 22 (20 - 25) | **<0.001** |
|  | 30-39 | 1330 | 238 | 18 (16 - 20) |  |
|  | 40-49 | 1414 | 208 | 15 (13 - 17) |  |
|  | 50-59 | 951 | 158 | 17 (14 - 19) |  |
|  | Over 60 | 282 | 45 | 16 (12 - 21) |  |
| **Sex** | Female | 3,959 | 669 | 17 (16 - 18) | **0.008** |
|  | Male | 1,126 | 229 | 20 (18 - 23) |  |
| **Ethnicity** | Irish | 3,798 | 595 | 16 (15 - 17) | **<0.001** |
|  | Any other white background | 476 | 94 | 20 (16 - 24) |  |
|  | Asian background | 599 | 148 | 25 (21 - 28) |  |
|  | African or any other black background | 117 | 39 | 33 (25 - 43) |  |
|  | Other | 95 | 22 | 23 (15 - 33) |  |
| **Country of birth** | Ireland | 3,630 | 567 | 16 (14 - 17) | **<0.001** |
|  | United Kingdom | 295 | 44 | 15 (11 - 20) |  |
|  | India | 293 | 76 | 26 (21 - 31) |  |
|  | Philippines | 214 | 54 | 25 (20 - 32) |  |
|  | Poland | 85 | 20 | 24 (15 - 34) |  |
|  | USA | 55 | 11 | 20 (10 - 33) |  |
|  | Romania | 45 | 15 | 33 (20 - 49) |  |
|  | Nigeria | 35 | 19 | 54 (31 - 71) |  |
|  | Other | 433 | 92 | 21 (17 - 25) |  |
| **Education** | Primary | 22 | 7 | 32 (14 - 55) | **<0.001** |
|  | Secondary | 609 | 133 | 22 (19 - 25) |  |
|  | Third level | 2,244 | 434 | 19 (18 - 21) |  |
|  | Post-graduate | 2,210 | 324 | 15 (13 - 16) |  |
| **Role** | Admin | 676 | 85 | 13 (10 - 15) | **<0.001** |
|  | Medical/dental | 713 | 116 | 16 (14 - 19) |  |
|  | Nursing/ midwifery | 1,891 | 395 | 21 (19 - 23) |  |
|  | Allied health | 1,044 | 118 | 11 (9.4 - 13) |  |
|  | General support | 365 | 81 | 22 (18 - 27) |  |
|  | Health care assistant | 291 | 93 | 32 (27 - 38) |  |
|  | Other | 105 | 10 | 10 (4.7 - 17) |  |
| **Lives with** | Alone | 464 | 61 | 13 (10 - 17) | **0.008** |
|  | With others | 4,610 | 834 | 18 (17 - 19) |  |
|  | Missing | 11 | 3 | 27 (6.0 - 61) |  |
| **Lives with HCW** | Yes | 1,571 | 340 | 22 (20 - 24) | **<0.001** |
|  | No | 3412 | 540 | 16 (16 - 17) |  |
|  | Missing | 102 | 18 | 18 (11 - 26) |  |

\*Calculated using the chi-square test

**Table 2b.**
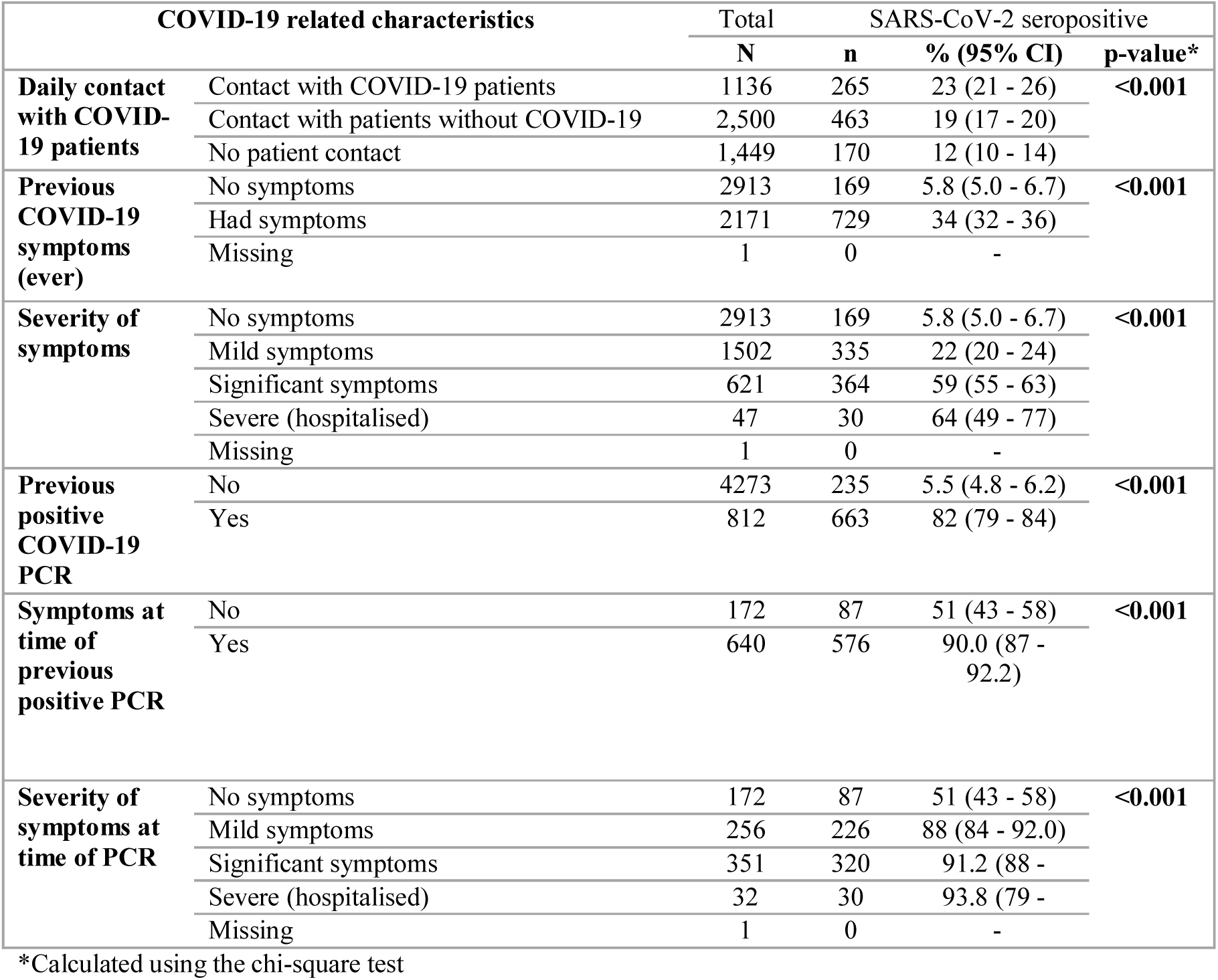
Prevalence of SARS-CoV-2 seropositivity by COVID-19 related characteristics, both hospitals, April 2021

#### SARS-CoV-2 seropositivity (due to natural infection) by previous diagnosis and symptoms

Sixteen percent (812/5085) of participants reported having had a PCR-confirmed infection with COVID-19 at some stage. Of these, 82% (663/812) were seropositive, and 21% (172/812) were asymptomatic at the time of their positive PCR, Table 2b. Seroprevalence among those that were symptomatic (576/640; 90%) was significantly higher than seroprevalence among those who were asymptomatic at the time of their confirmed COVID-19 infection (87/172; 51%; p<.001). Nineteen percent (169/898) of those with detectable antibodies had never experienced symptoms consistent with COVID-19 infection.

#### Undiagnosed SARS-CoV-2 infection

In total, 898 participants were seropositive (due to natural infection). Of these, 235/898 (26%) had never been diagnosed with COVID-19 infection, representing 4.6% (235/5085) of the total study population. Just over half of those with undiagnosed infection (121/235; 51%) had experienced COVID-19 like symptoms at some stage; of those 74% (89/121) had experienced mild symptoms and 26% (32/121) had experienced moderate symptoms. This proportion of symptomatic undiagnosed participants who experienced only mild symptoms (89/121, 74%) was much higher than the proportion of symptomatic diagnosed participants who experienced only mild symptoms (226/576; 39%) (Table A, Annex, main report, (31)). Most participants with undiagnosed infection reported daily patient contact in their role (192/235; 82%). Detailed analysis of undiagnosed infection by HCW role and by hospital is shown in Table 2i, Annex.

#### Risk factors for SARS-CoV-2 antibody positivity

Characteristics of those participants who were seropositive compared with those who were seronegative for both hospitals combined are shown in Tables 2a and 2b. On multivariable analysis of the combined hospital data, the adjusted relative risk (aRR) of seropositivity was statistically significant for the following characteristics: working in Hospital 1 (aRR 1.5, 95% CI 1.3-1.8, p<.001), being a healthcare assistant (aRR 1.8, 95% CI 1.3-2.3, p<.001), being of African or other black background (aRR 1.7, 95% CI 1.3-2.2, p<.001), lower level of education (aRR 1.4 for secondary level education, 95% CI 1.1-1.8, p=0.002), being a nurse (aRR 1.4, 95% CI 1.0-1.8, p=0.022), daily contact with COVID-19 patients (aRR 1.4, 95% CI 1.1-1.7, p=0.002), daily contact with patients without suspected or confirmed COVID-19 (aRR 1.3, 95% CI 1.1-1.5, p=0.013), being 18-29 years of age (aRR 1.3, 95% CI 1.1-1.6, p=0.002), being male (aRR 1.2, 95% CI 1.0-1.4, p=0.016), and living with other HCW (aRR 1.2, 95% CI 1.0-1.4, p=0.007), Table 3a. Multivariable analysis by hospital is shown in Tables 3b and 3c, Annex.

**Table 3a.**
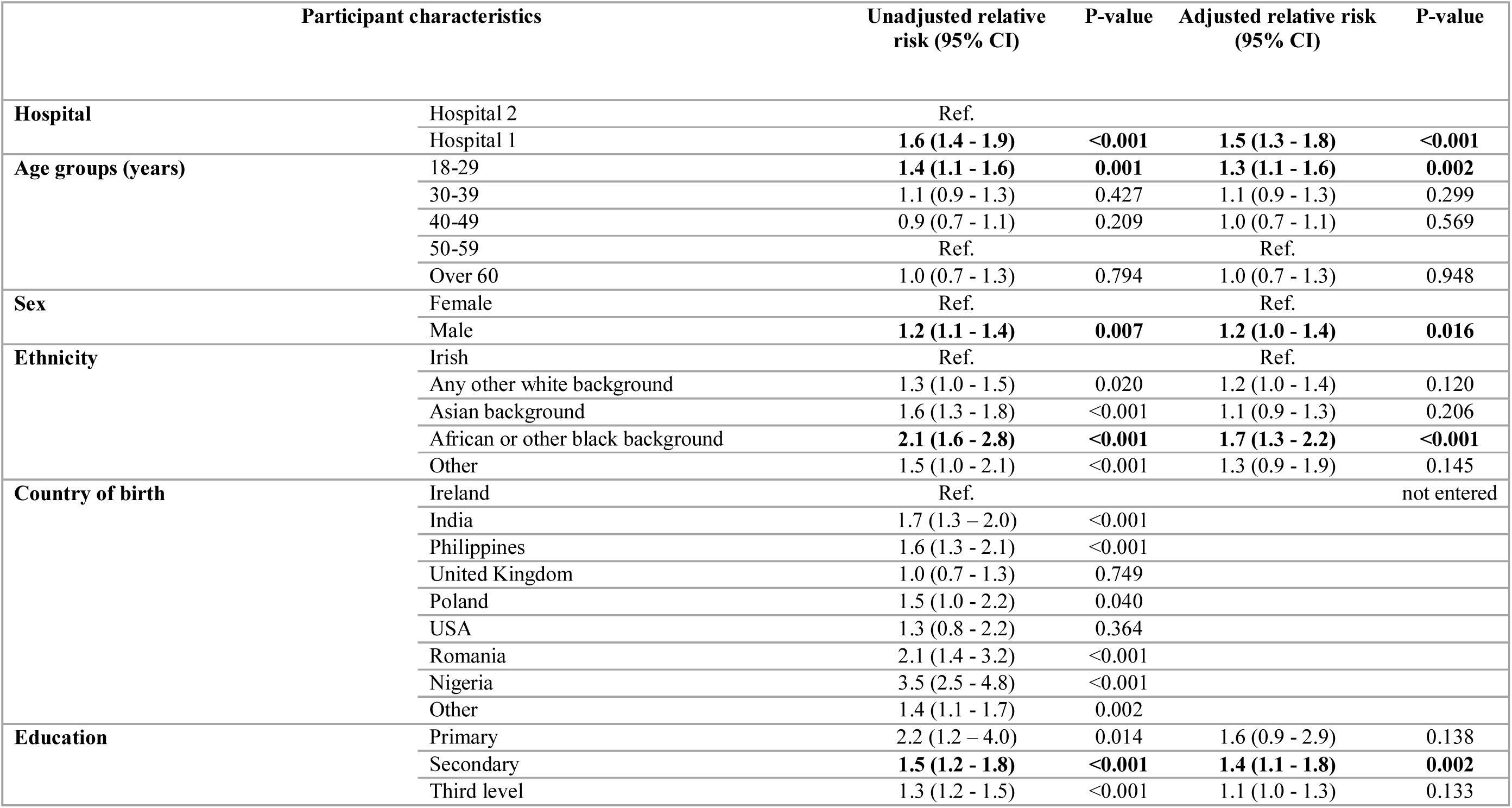
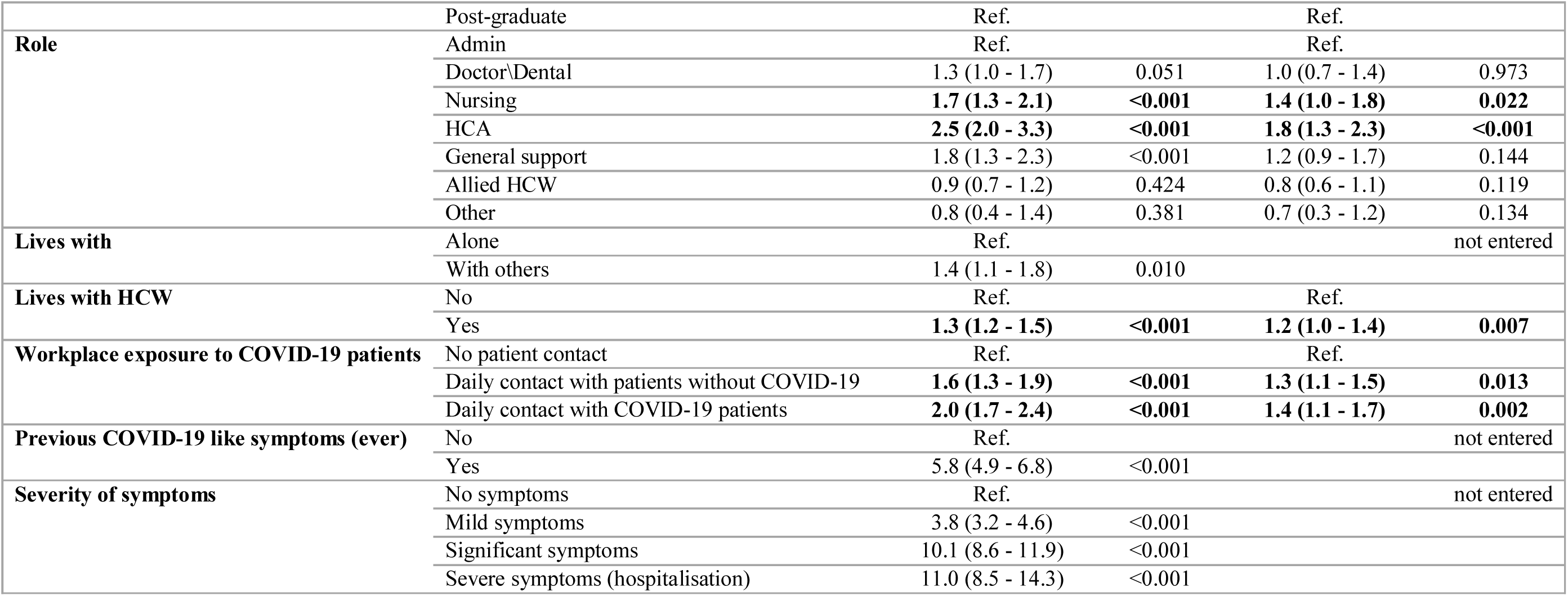
Association between risk factors and SARS-CoV-2 seropositivity, both hospitals, April 2021

### 2. Antibody response to vaccination

In total, 80% (4111/5085) of participants were fully vaccinated and a further 14% (716/5085) were partially vaccinated. All fully vaccinated participants had received the Pfizer vaccine and the majority of partially vaccinated participants (681/716, 95%) had received the Vaxzevria vaccine (due to timing of vaccine roll-out (which commenced with the Pfizer vaccine) and due to the longer dosing interval of the Vaxzevria vaccine (28)). Vaccination uptake by ethnicity was similar, with 142/3661 (3.9%, 95% CI 3.3-4.6)) of White Irish participants being unvaccinated, compared to 30/571 (5.2%, 95% CI 3.7-7.4) of Asian participants and 7/110 (6.4%, 95% CI 3.1-13) of Black participants. All fully vaccinated participants (4111/4111, 100%) and 99.6% (713/716) of partially vaccinated participants had detectable anti-S antibodies.

### 3. SARS-CoV-2 infection post vaccination

In total, 116 participants reported PCR-confirmed SARS-CoV-2 infection since vaccination; of those 23/116 (20%) were fully vaccinated, representing 0.6% (23/4111) of all fully vaccinated participants. There were 93 infections in partially vaccinated participants representing 93/716 (13%; 95% CI 11 - 16) of partially vaccinated participants having had a PCR-confirmed infection post vaccination compared to 23/4111 (0.6%; 95% CI 0.4 - 0.8) of fully vaccinated participants (p-value= <.001 (chi-square)). All fully and partially vaccinated participants with infection post-vaccination had anti-spike antibodies detected.

Of the 23 breakthrough infections in fully vaccinated participants, all had received the Pfizer vaccine. (It is noted that recipients of other vaccines in our study were not yet fully vaccinated). The median interval between first and second vaccine dose was 21 days (as recommended before 18^th^ January 2021). For those 23 participants, the median number of days between second vaccine dose and PCR positive test was 30 days (IQR 25-50 days). Five (22%) had symptoms at the time of the positive PCR test and 18 (78%) did not have symptoms. Characteristics of participants with breakthrough infection are shown in Table F annex.

#### Comparison of crude seroprevalence from October 2020 to April 2021

In Hospital 1, the seroprevalence significantly increased from 15% in October 2020 to 21% in April 2021 (p<.001), and in Hospital 2 the seroprevalence significantly increased from 4.1% in October 2020 to 13% in April 2021 (p<.001). For both hospitals combined, seroprevalence increased from 10% in October 2020 to 18% in April 2021. By ethnic group, the increase in seroprevalence was most pronounced for HCWs of African or other black background (from 14% in October 2020 to 33% in April 2021; p=0.001). By education level, increase in seroprevalence was highest for HCWs with lower education levels; the increase for those of primary education (from 14% in October 2020 to 32% in April 2021) was high but not significant due to low numbers in this subgroup (p=0.121); for those of secondary level education the increase was also high (from 8.9% to 22%) and it was significant (p<0.001). By professional subgroup, increase in seroprevalence was highest for general support staff (from 7.6% in October 2020 to 22% in April 2021; p<0.001) and for HCAs (from 18% in October 2020 to 32% in October 2021; p<0.001). A comparison of crude seroprevalence in October 2020 and in April 2021 by participant characteristics and by hospital is shown in Table 4, Annex.

## Discussion

### Participation and demographics

Our participants were similar in age and sex to those in other European studies (32) (33) (7). The participation rate was acceptable for an institutional opt-in study, was comparable to other European studies (34), and included representation from all HCW groups, including the traditionally harder-to-reach groups such as general support staff and healthcare assistants, who may not engage as frequently with hospital communications platforms.

### Overall seroprevalence

The difference in seroprevalence between the two sites primarily reflects the difference in community incidence, with a corresponding higher seroprevalence seen in the more densely populated capital city of Dublin, which has had higher community incidence throughout the pandemic thus far. Other studies have also shown community incidence to be one of the main factors impacting risk to HCW (35) (36). The rise in seroprevalence at both sites reflects the magnitude of the third wave of the pandemic throughout Ireland. The relatively higher increase in seroprevalence in Hospital 2 compared to Hospital 1 likely reflects the fact that the community incidence in the Galway area approached that of the Dublin incidence during this third COVID-19 wave (25).

The difference in overall seroprevalence was also likely to have been impacted by differences in hospital infrastructure, work-practices, bed-flow management, and the differing demographic and social factors by HCW role at each site. Broad work-place practices in both hospitals have been similar throughout the pandemic, including ward-based medical teams and universal use of face masks. There were no issues with personal protective equipment (PPE) availability in either of the hospitals involved in our study at any stage thus far during the pandemic, and where staff were re-deployed to improve the hospital’s capacity to deal with the outbreak, staff were not deployed to areas that would have been outside of their scope of practice, and all staff had training on the correct use of PPE. Both hospitals experienced multiple outbreaks during the 3^rd^ wave of the pandemic, however the absence of whole genomic sequencing (WGS) in this study precludes identifying the role of hospital outbreaks in influencing the overall seroprevalence. A recently published Dutch study (conducted prior to HCW vaccination) used WGS to show that transmission to HCW was largely from other HCW – interestingly, they showed a complete absence of transmission from patients to HCW (37). While this may be the case in our cohort, the fact that HCW role and patient proximity were independent risk factors for seropositivity lead us to believe that acquisition by HCW in the hospital setting may have been from both patients and other HCW. It is also difficult to identify the proportion of the risk that is attributable to the workplace versus the community/ household/ social factors. It is likely that transmission also took place in the household, where higher attack rates are seen (38). It is also unclear if differential timing and spread of VOCs in each location may have impacted overall seroprevalence. Accurate assessment of the risk to HCW in each of these situations and environments, and the impact of VOCs on overall seroprevalence, would necessitate WGS of all HCW with confirmed infection. Real time results would be ideal in order to inform local IPC measures.

The overall seroprevalence in both hospitals was higher than the European average of 8.5% in a recently published meta-analysis (39), however this meta-analysis only took into account studies published up until August 2020. Individual studies across Europe in the first year of the pandemic showed a wide range of SARS-CoV-2 seroprevalence among HCW (1-45%) (32) (7) (40) (41) (42) (43). A recently published study of over 80,00 HCW in Italy showed a seroprevalence of 12% amongst HCW, however the serological testing was conducted in April and May 2020 (34). Assumably the seroprevalence amongst HCW has risen across the rest of Europe and worldwide since then, as our study shows it has in Ireland, however, to the best of our knowledge, there are no published studies estimating SARS-CoV-2 seroprevalence in HCW in Europe in 2021, nor comparing this to seroprevalence in 2020.

### Seroprevalence by role and type of patient contact

Daily contact with patients with known/suspected COVID-19 infection was associated with higher seroprevalence, followed by daily contact with patients without known or suspected COVID-19 infection. HCWs with little or no patient contact had the lowest seroprevalence, however we showed in October 2020 that non-patient-facing staff, although less likely to be seropositive, were more likely to have had an undiagnosed infection (i.e. to be antibody positive with no history of PCR-confirmed COVID-19 infection (31)). This reflects the findings of other studies, including our own findings in these same hospitals in October 2020 (24) (44) (45) (46). In terms of working role, being a nurse or a HCA carried a higher aRR, likely also reflecting the close patient contact involved in performing these roles. This finding has also been shown in other large European studies (34).

The seroprevalence among general support staff (which includes domestic and catering staff, maintenance, security and porters) trebled from October 2020 to April 2021. This increase was in both locations and was across all groups in this category (Table 2f, Annex). This was possibly related to outbreaks amongst these staff groups, though it was not clear whether these outbreaks were related to the workplace or not. There may be improper compliance with use of PPE, and fatigue with other elements of infection prevention and control (IPC) precautions as the pandemic has progressed. There are likely to also be other social factors involved that our study was not designed to assess.

### Previous symptoms and testing

Nineteen percent of participants with detectable antibodies reported never having experienced symptoms that were consistent with infection with COVID-19. This falls within the broad range reported by other studies (47). Those who had symptomatic infection had a higher rate of antibody positivity than those who had an asymptomatic infection (90% versus 51%), which is also in keeping with other published data (48). Over a quarter of participants reported having COVID-19 like symptoms at some stage but never having a positive PCR. This highlights the potential overlap in symptoms with other circulating viruses, including rhinoviruses which were circulating widely over the winter of 2020/21 in Ireland (49), and is a reminder of the impossibility of clinically excluding COVID-19 infection in HCW with symptoms, including in those with only mild symptoms, especially over the winter period. It also highlights the complexity involved in developing case definitions and testing guidelines for symptomatic individuals. A subset of these participants never sought testing, despite their symptoms, which reinforces the need for clear messaging about availability of and indication for testing.

### Undiagnosed infections

In both hospitals, the seroprevalence was higher than the known PCR-confirmed diagnoses of COVID-19 infection of the same timeframe (21% vs 18% in Hospital 1, and 13% vs 9.2% in Hospital 2) (50). Over a quarter of infections were undiagnosed, and those with an undiagnosed infection were more likely to have been asymptomatic. This proportion of undiagnosed infection had decreased from 39% in October 2020, assumedly due to increased testing and increased awareness, however it remains high despite both hospitals having onsite PCR testing available to HCW with symptoms or those identified as a close contact with a confirmed case of COVID-19 from mid-March 2020. Those with undiagnosed infection fall into two categories; those with asymptomatic infection, and those who did have symptoms but did not seek testing. Most of these undiagnosed infections were associated with none or only mild symptoms, however it is still possible that these undiagnosed HCW were working during the infectious period, with potential for onwards transmission to patients and other staff members if proper use of PPE and other IPC measures were not strictly adhered to.

Onwards transmission in the household is even more likely in this setting, where ongoing unprotected close contact is likely. Easy access to testing, early detection of infection, and ongoing adherence to standard infection control precautions at all times, as well as the appropriate use of PPE including face masks in the hospital setting, irrespective of symptoms remain important (51). This finding also supports the recommendation for screening of asymptomatic staff, including vaccinated staff in certain situations, when a hospital outbreak of infection with COVID-19 occurs. The exact role and methodology of routine asymptomatic screening, either widespread or in certain HCW groups, remains to be defined. Regarding those who had symptoms but did not seek testing, concerted efforts need to be made to ensure all HCW groups, including those who are non-patient-facing, are aware of the availability of and need to seek advice regarding testing.

### Risk factors for antibody positivity

The main risk factors statistically significantly associated with antibody positivity (in decreasing order of aRR) were being a HCA, being of Black ethnicity, working in Hospital 1, lower level of education, being a nurse, having daily contact with patients (especially those known or suspected to have COVID-19 infection), being age 18-29 years, living with other HCW and being male. Seroprevalence by age and sex were similar to previously published literature (6) (24) (39). Similar findings of increased risk with direct patient contact, the role of HCA, and working with patients with COVID-19 infection have also been reported in the literature (7) (39) (52) (53).

Apart from the changes in seroprevalence by role discussed above, there were two main new findings on multivariable analysis between October 2020 and April 2021. Firstly, lower level of education emerged as an independent risk factor for seropositivity. Lower socio-economic status has been previously noted to correlate with increased risk of COVID-19 infection, and increased risk of poor clinical outcomes (54) (55). The second notable new finding was a change in the seropositivity by ethnic group; the seroprevalence amongst those of Black ethnicity trebled (from 14% (16/113) to 43% (39/117), for an aRR of 1.7 (95% CI 1.3-2.2, p<.001)). They were also more likely to be asymptomatic, but not more likely to have an undiagnosed infection. Those of Asian ethnicity had a higher risk of seropositivity in October 2020, but this finding was no longer significant in April 2021. Both of these ethnic groups, as well as other minority ethnic groups which were likely under-represented in our study, have been shown to have increased risk in other studies (39) (56) (57) (58). There are likely to be social factors contributing to these ethnicity-related findings in both hospitals that our study did not measure. Ethnicity and level of education have been shown throughout the pandemic to be associated with seropositivity and with poor outcomes of COVID-19 infection. There may be some unmodifiable factors contributing to this risk, however all efforts need to continue to be made to mitigate modifiable risk factors amongst these groups; we cannot assume, even at this stage of the pandemic, that educational messaging regarding risk reduction has reached all groups equally.

Living with other HCW carried an increased risk for seropositivity, similar to our previous findings. This supports the theory that a proportion of the HCW contracting COVID-19 are doing so in their home environment. This finding was stronger in Hospital 1, where the community incidence was higher and the density of shared living space is also likely to be higher. Other studies have found correlation between size of household and antibody positivity (32) as well as higher risk of COVID-19 with a known household contact (59). To the best of our knowledge ours is the first study to comment on a significant risk of antibody positivity in HCW living with other HCW. This finding was common to both of our serosurveys (24).

### Antibody response to vaccination

Most participants were either fully or partially vaccinated. All fully vaccinated participants, and the majority of partially vaccinated participants, had detectable anti-S antibodies. Other studies have also shown high rates of seropositivity after both first and second dose vaccination (27) (60) (61). There were less infections in those fully vaccinated than in those partially vaccinated (13% versus 0.6% of participants reported infection post first and second vaccine dose respectively), despite the fact that almost all of these participants had detectable antibodies. The 23 breakthrough infections, in 0.6% of the fully vaccinated study population, are in keeping with the rate of breakthrough infections experienced internationally (62). These breakthrough infections serve as a reminder that vaccination does not prevent infection acquisition, even in the setting of confirmed serological response to vaccination. Most of those with breakthrough infections had no symptoms, in keeping with the literature on vaccine effectiveness in reduction of severe symptoms and hospitalisation (15) (63) however the numbers are too small for any statistical comparison with symptoms in those who were unvaccinated. It would be prudent for all IPC measures to remain in place in the hospital setting, including for vaccinated HCW, while research is ongoing into the effects of vaccination on infection acquisition and onwards transmission of SARS-CoV-2, including with VOCs. Vaccinated HCW should not be exempt from measures discussed above in relation to minimising the rate of undiagnosed infections (access to testing, adherence to standard IPC precautions and inclusion in screening of asymptomatic staff in the case of a hospital outbreak).

## Limitations

This study has several limitations. Firstly, information on COVID-19 symptoms and PCR test results were self-reported and thus could be biased and could not be verified. Secondly, although the uptake rate was good for an opt-in study, it was lower than the uptake for PRECISE 1; declining interest in research in the area of COVID-19 is a natural phenomenon as the pandemic progresses. Many staff at this point in time already knew they had been infected and therefore may have less interest in participating and availing of serology testing. Most staff had been vaccinated and therefore may also have a degree of comfort that produces less interest in knowing their antibody status. This could lead to underestimation of the overall seroprevalence. PRECISE 2 took place during the third wave of the pandemic in Ireland, which was the largest in magnitude and the longest. Other limitations include that WGS data were not available, particularly for those infected after full vaccination, and also that information on biological factors, e.g. co-morbidities, was not available. Although the communication strategy was an important part of the recruitment process, the study took part during our third wave of the pandemic, during Level 5 restrictions - the highest level of COVID-19 national restrictions - and therefore also relied heavily on engagement with information technology (IT) platforms (email, messenger groups, hospital intranet) and less on face-to-face announcements. Thirdly, as with all opt-in studies, there may be a selection bias. Those who had been vaccinated may have had less interest in participating due to less curiosity about their own serostatus, and therefore we may have underestimated the overall vaccination status of the workforce (however vaccination coverage amongst our study participants equates to local unpublished hospital data so we feel the study population was representative on this front). Conversely, those who were unvaccinated may have had a fear of having to announce their vaccine-status to the study team, despite results not being linked to occupational health records, and those who had been vaccinated may have been more likely to participate as they may be more likely to have health seeking behavior. The online consent process, questionnaire, and blood test booking system risks exclusion of those who are less literate in IT. This was identified as a potential limitation from the start, and attempts were made to mitigate this selection bias. Multilanguage information and plain English were used, and groups identified as potentially at risk of exclusion on this basis were targeted directly for inclusion in the study, with small-group sessions to aid consent and questionnaire completion and walk-in clinics for phlebotomy. We do not have individual level information on reasons for non-participation, or socio-demographic status of non-participants for comparison, but level of uptake by professional role was deemed to be representative of the hospital HCW population in both hospitals. The absence of genomic sequencing data precludes identifying the role of hospital outbreaks in influencing the overall seroprevalence, as well as drawing any definite conclusions regarding attributable risk to the workplace versus the community for HCW.

Detection of anti-spike antibody in conjunction with a self-reported history of vaccination was considered to be as a response to vaccination. It is possible that some of these participants may have detectable anti-spike antibody related to natural infection; these were not counted when assessing overall seroprevalence of presumed past infection, and therefore this overall seroprevalence may be an underestimate. Seroprevalence could also be underestimated because recent infection could be missed as several days are required for seroconversion of SARS-CoV-2 infection. Finally, some false negatives and false positives are expected with all laboratory tests. When the estimate of seroprevalence is adjusted using the manufacturer’s stated specificity of 99.8% and sensitivity of 99.5%, the seroprevalence does not change (18%; 95% CI 17% - 19%).

## Conclusion and Recommendations

To the best of our knowledge there is no other published study to date estimating SARS-CoV-2 seroprevalence in a large HCW group in 2021. This study is a unique comparison between two hospitals in areas of differing community incidence over time, and at two points in time. The rise in seroprevalence between October 2020 and April 2021 reflects the magnitude of the third wave of the pandemic in both locations, and the difference in community incidence in each location. Many of the risk factors identified point towards hospital acquisition (HCW role and type of patient contact) while others point towards household acquisition (living with others, living with other HCW); genomic sequencing is needed to identify the role of hospital outbreaks in influencing the overall seroprevalence, as well as to draw conclusions regarding attributable risk to the healthcare environment versus the community or household.

Our study highlights the changing epidemiology with different waves of the pandemic, and in different locations. More serological studies are needed in HCW worldwide to assess these changes in risk factors as the pandemic progresses. While each wave of the pandemic may be associated with different risk factors in different locations, many risk factors also remain constant as the pandemic continues worldwide; the association of seropositivity with lower level of education and ethnicity a year and a half into the pandemic may indicate that some groups have not been adequately reached by messaging and education, and concerted efforts are needed to specifically mitigate modifiable risk factors due to ethnicity and level of education.

The antibody response to vaccination is reassuring however further studies are needed to correlate serological response with functional immunity and to estimate duration of protection from infection, particularly with the ongoing emergence of variants of concern. With emerging evidence of reduction in transmission from vaccinated individuals, the authors strongly endorse immediate vaccination of all HCW who have not yet been vaccinated. We did, however, show confirmed infection in a small minority of fully vaccinated participants, all with appropriate antibody response to the vaccine, therefore messaging to HCW regarding the role and limits of vaccination need to be clear and should include the ongoing risk of infection and transmission.

The proportion of infections that were previously undiagnosed decreased from October 2020 to April 2021 but remained high. In light of both the undiagnosed infections and the breakthrough infections in vaccinated individuals, ongoing adherence to all infection prevention and control standards in the healthcare setting and household are paramount. It is essential that HCW have easy access to testing, even with mild or no symptoms, and even in the post vaccination setting (64). We recommend ongoing risk assessment in the setting of a hospital outbreak, and where indicated, screening of HCW, including those without symptoms, and including those who are vaccinated. Breakthrough infections will continue to occur in fully vaccinated individuals and need to be further characterised; HCW are an ideally positioned group for this due to their increased exposure, and in the interim, we should not change established infection prevention and control measures or testing protocols amongst HCW on the basis of vaccination alone. Assessment of vaccinated HCW with PCR-confirmed SARS-CoV-2 infection should include SARS-CoV-2 WGS and, if possible, SARS-CoV-2 WGS of index cases identified by follow-up, as well as assessment of potential host determinants of infection, in order to advance understanding of risk factors for breakthrough infection.

## Data Availability

All data produced in the present study are available upon reasonable request to the authors

## Acknowledgements

We would like to acknowledge the National Public Health Emergency Team (NPHET) who supported this study, the study steering group who planned the study and critically evaluated this manuscript, the study team who coordinated the running of the study in each hospital, the hospital management at both sites for their support for the study, and the staff of both hospitals who participated. We would especially like to acknowledge the phlebotomy departments in each hospital for facilitating the sampling of over 5000 participants, the virology and biochemistry laboratories in each hospital for processing the samples and the human resources department for their help with denominator data. This work was supported financially by the Irish Health Service Executive COVID-19 budget.

## Authors’ Contributions

NA was the Principal Investigator for the study, designed and executed the study in collaboration with the lead site investigators and the Study steering group. NA organised the data collection, aided interpretation of the data and wrote the scientific manuscript. MB analysed the data with the support of AICM, LD and CW. CB and CF were the lead site investigators and aided with the design and execution of all aspects of the study. NC and UNR coordinated the laboratory testing of all samples. LD was the study coordinator and oversaw all aspects of the running of the study, and coordinated the PRECISE Study Steering Group, who aided with study design and execution. EH and CK supported data collection. All named authors reviewed the final manuscript.

## Data Availability Statement

This dataset is not available to the public for ethical reasons of data protection as certain individuals may be identifiable from the data.

## Annex

**Table A.**
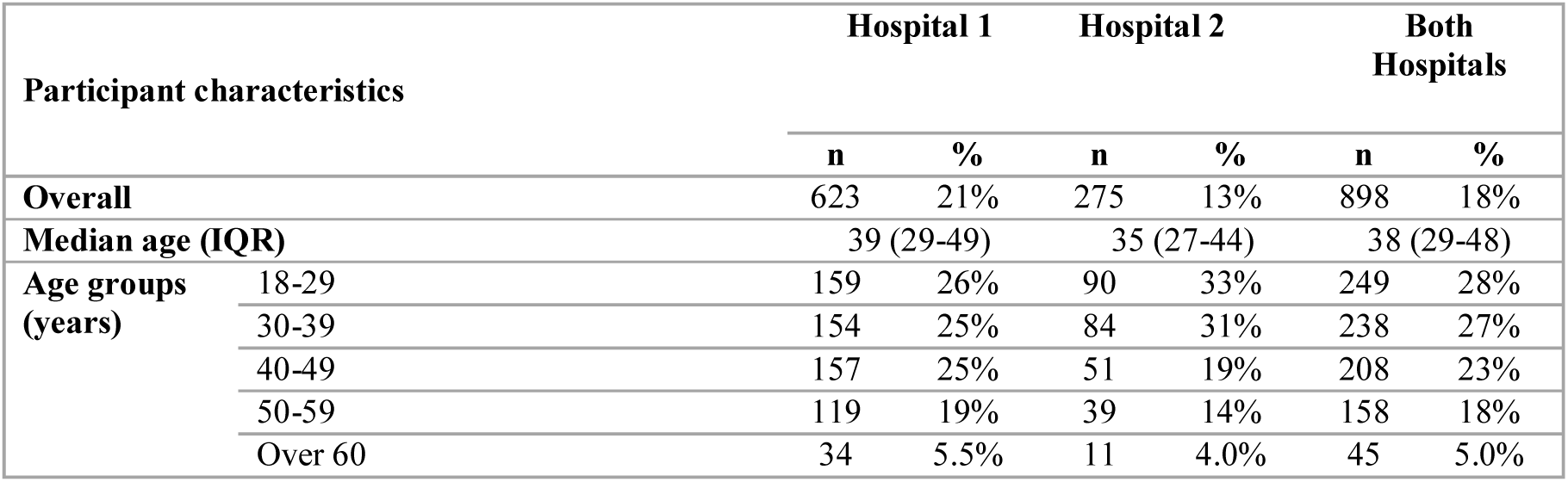
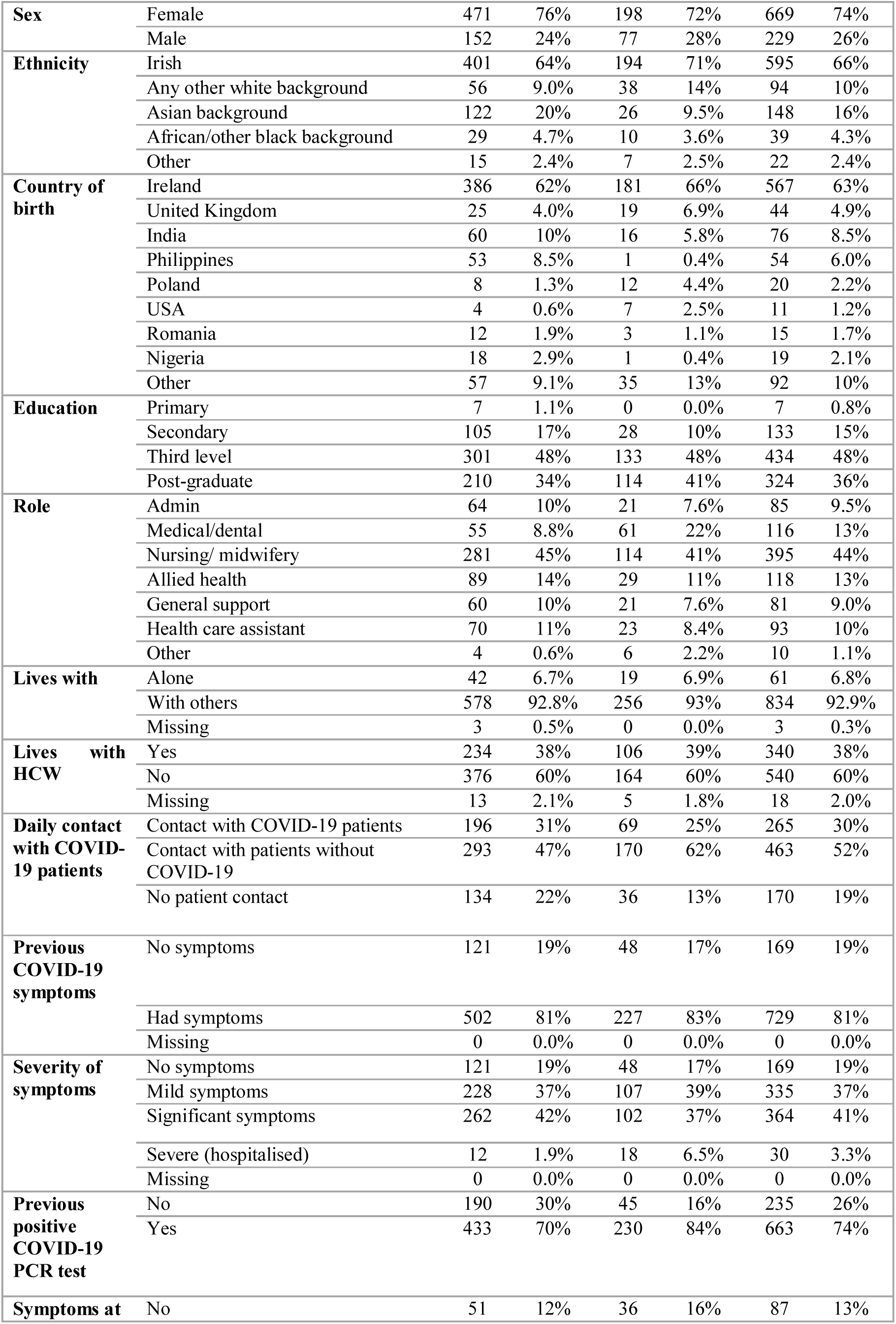
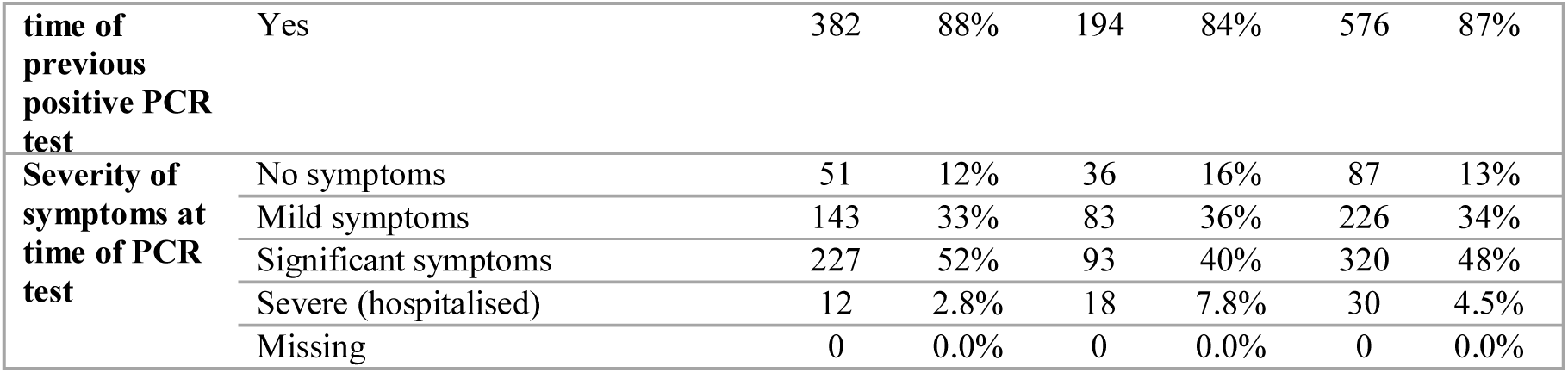
Characteristics of HCWs with SARS-CoV-2 seropositivity (n=898), by hospital, April 2021

**Table 2c.** Prevalence of SARS-CoV-2 seropositivity by participant characteristics, Hospital 1, April 2021

| Participant characteristics |  | Total | SARS-CoV-2 seropositive |  |  |
| --- | --- | --- | --- | --- | --- |
|  |  | N | n | % (95% CI) | p-value* |
| **Overall** |  | 2945 | 623 | 21 (20 - 23) | - |
| **Age groups (years)** | 18-29 | 653 | 159 | 24 (21 - 28) | 0.184 |
|  | 30-39 | 765 | 154 | 20 (17 - 23) |  |
|  | 40-49 | 811 | 157 | 19 (17 - 22) |  |
|  | 50-59 | 565 | 119 | 21 (18 - 25) |  |
|  | Over 60 | 151 | 34 | 23 (16 - 30) |  |
| **Sex** | Female | 2,278 | 471 | 21 (19 - 22) | 0.240 |
|  | Male | 667 | 152 | 23 (20 - 26) |  |
| **Ethnicity** | Irish | 2,091 | 401 | 19 (18 - 21) | <0.001 |
|  | Any other white background | 257 | 56 | 22 (17 - 27) |  |
|  | Asian background | 470 | 122 | 26 (22 - 30) |  |
|  | African or any other black background | 69 | 29 | 42 (30 - 55) |  |
|  | Other | 58 | 15 | 26 (15 - 39) |  |
| **Country of birth** | Ireland | 2,025 | 386 | 19 (17 - 21) | 0.001 |
|  | United Kingdom | 134 | 25 | 19 (12 - 26) |  |
|  | India | 225 | 60 | 27 (21 - 33) |  |
|  | Philippines | 198 | 53 | 27 (21 - 34) |  |
|  | Poland | 26 | 8 | 31 (14 - 52) |  |
|  | USA | 21 | 4 | 19 (5.4 - 42) |  |
|  | Romania | 40 | 12 | 30 (17 - 47) |  |
|  | Nigeria | 25 | 18 | 72 (51 - 88) |  |
|  | Other | 251 | 57 | 23 (18 - 28) |  |
| **Education** | Primary | 20 | 7 | 35 (15 - 59) | <0.001 |
|  | Secondary | 409 | 105 | 26 (22 - 30) |  |
|  | Third level | 1,280 | 301 | 24 (21 - 26) |  |
|  | Post-graduate | 1,236 | 210 | 17 (15 - 19) |  |
| **Role** | Admin | 403 | 64 | 16 (12 - 20) | <0.001 |
|  | Medical/dental | 357 | 55 | 15 (12 - 20) |  |
|  | Nursing/ midwifery | 1097 | 281 | 26 (23 - 28) |  |
|  | Allied health | 612 | 89 | 15 (12 - 18) |  |
|  | General support | 243 | 60 | 25 (19 - 31) |  |
|  | Health care assistant | 179 | 70 | 39 (32 - 47) |  |
|  | Other | 54 | 4 | 7.4 (2.1 - 18) |  |
| **Lives with** | Alone | 270 | 42 | 16 (11 - 20) | 0.019 |
|  | With others | 2,667 | 578 | 22 (20 - 23) |  |
|  | Missing | 8 | 3 | 38 (8.5 - 76) |  |
| **Lives with HCW** | Yes | 928 | 234 | 25 (22 - 28) | <.001 |
|  | No | 1,964 | 376 | 19 (17 - 21) |  |
|  | Missing | 53 | 13 | 25 (14 - 38) |  |

\*Calculated using the Chi-square test

**Table 2d.** Prevalence of SARS-CoV-2 seropositivity by COVID-19 related characteristics, Hospital 1, April 2021

| COVID-19 related characteristics |  | Total | SARS-CoV-2 seropositive |  |  |
| --- | --- | --- | --- | --- | --- |
|  |  | N | n | % (95% CI) | p-value* |
| **Daily contact with COVID-19 patients** | Contact with COVID-19 patients | 726 | 196 | 27 (24 - 30) | <0.001 |
|  | Contact with patients without COVID-19 | 1,362 | 293 | 22 (19 - 24) |  |
|  | No patient contact | 857 | 134 | 16 (13 - 18) |  |
| **Previous COVID-19 symptoms** | No symptoms | 1571 | 121 | 7.7 (6.4 - 9.1) | <0.001 |
|  | Had symptoms | 1374 | 502 | 37 (34 - 39) |  |
|  | Missing | 0 | 0 | - |  |
| **Severity of symptoms** | No symptoms | 1571 | 121 | 7.7 (6.4 - 9.1) | <0.001 |
|  | Mild symptoms | 932 | 228 | 24 (22 - 27) |  |
|  | Significant symptoms | 420 | 262 | 62 (58 - 67) |  |
|  | Severe (hospitalised) | 21 | 12 | 57 (34 - 78) |  |
|  | Missing | 1 | 0 | - |  |
| **Previous positive COVID-19 PCR test** | No | 2427 | 190 | 7.8 (6.8 - 9.0) | <0.001 |
|  | Yes | 518 | 433 | 84 (80 - 87) |  |
| **Symptoms at time of previous positive PCR** | No | 95 | 51 | 54 (43 - 64) | <0.001 |
|  | Yes | 423 | 382 | 90.3 (87 - 93.0) |  |
| **Severity of symptoms at time of PCR test** | No symptoms | 95 | 51 | 54 (43 - 64) | <0.001 |
|  | Mild symptoms | 162 | 143 | 88 (82 - 92.8) |  |
|  | Significant symptoms | 248 | 227 | 91.5 (87 - 94.7) |  |
|  | Severe (hospitalised) | 12 | 12 | 100 (74 - 100) |  |
|  | Missing | 1 | 0 | - |  |

\*Calculated using the Chi-square test

**Table 2e.** Prevalence of SARS-CoV-2 seropositivity by participant characteristics, Hospital 2, April 2021

| Participant characteristics |  | Total | SARS-CoV-2 seropositive |  |  |
| --- | --- | --- | --- | --- | --- |
|  |  | N | n | % (95% CI) | p-value* |
| **Overall** |  | 2140 | 275 | 13 (11 - 14) | - |
| **Age groups (years)** | 18-29 | 455 | 90 | 20 (16 - 24) | <0.001 |
|  | 30-39 | 565 | 84 | 15 (12 - 18) |  |
|  | 40-49 | 603 | 51 | 8.5 (6.4 - 11) |  |
|  | 50-59 | 386 | 39 | 10 (7.3 - 14) |  |
|  | Over 60 | 131 | 11 | 8.4 (4.3 - 15) |  |
| **Sex** | Female | 1,681 | 198 | 12 (10 - 13) | 0.005 |
|  | Male | 459 | 77 | 17 (13 - 21) |  |
| **Ethnicity** | Irish | 1,707 | 194 | 11 (10 - 13) | 0.002 |
|  | Any other white background | 219 | 38 | 17 (13 - 23) |  |
|  | Asian background | 129 | 26 | 20 (14 - 28) |  |
|  | African or any other black background | 48 | 10 | 21 (10 - 35) |  |
|  | Other | 37 | 7 | 19 (8.0 - 35) |  |
| **Country of birth** | Ireland | 1605 | 181 | 11 (10 - 13) | <0.001 |
|  | United Kingdom | 161 | 19 | 12 (7.3 - 18) |  |
|  | India | 68 | 16 | 24 (14 - 35) |  |
|  | Poland | 59 | 12 | 20 (11 - 33) |  |
|  | USA | 34 | 7 | 21 (8.7 - 38) |  |
|  | Philippines | 16 | 1 | 6.3 (0.2 - 30) |  |
|  | Nigeria | 10 | 1 | 10 (0.3 - 45) |  |
|  | Romania | 5 | 3 | 60 (14 - 95) |  |
|  | Other | 182 | 39 | 19 (13 - 36) |  |
| **Education** | Primary | 2 | 0 | - | 0.485 |
|  | Secondary | 200 | 28 | 14 (10 - 20) |  |
|  | Third level | 964 | 133 | 14 (12 - 16) |  |
|  | Post-graduate | 974 | 114 | 12 (10 - 14) |  |
| **Role** | Admin | 273 | 21 | 7.7 (4.8 - 12) | <0.001 |
|  | Medical/dental | 356 | 61 | 17 (13 - 21) |  |
|  | Nursing/ midwifery | 794 | 114 | 14 (12 - 17) |  |
|  | Allied health | 432 | 29 | 6.7 (4.5 - 9.5) |  |
|  | General support | 122 | 21 | 17 (11 - 25) |  |
|  | Health care assistant | 112 | 23 | 21 (13 - 29) |  |
|  | Other | 51 | 6 | 12 (4.4 - 24) |  |
| **Lives with** | Alone | 194 | 19 | 10 (6.0 - 15) | 0.180 |
|  | With others | 1,943 | 256 | 13 (12 - 15) |  |
|  | Missing | 3 | 0 | - |  |
| **Lives with HCW** | Yes | 643 | 106 | 16 (14 - 20) | 0.001 |
|  | No | 1448 | 164 | 11 (10 - 13) |  |
|  | Missing | 49 | 5 | 10 (3.4 - 22) |  |

\*Calculated using the Chi-square test

**Table 2f.**
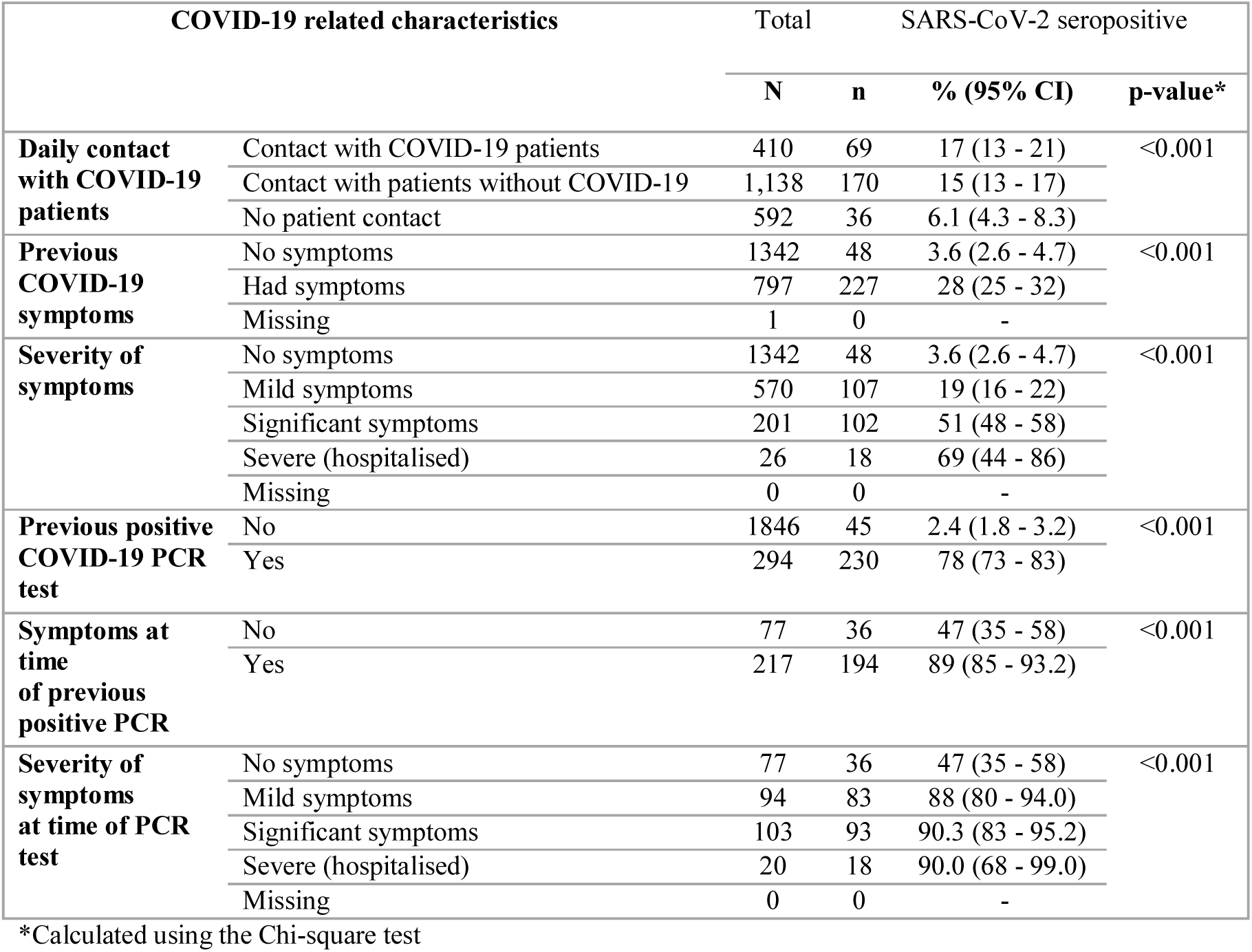
Prevalence of SARS-CoV-2 seropositivity by COVID-19 characteristics, Hospital 2, April 2021

**Table 2g.** Prevalence of SARS-CoV-2 seropositivity for general support staff, by role and hospital, PRECISE 2, April 2021

| Role | Hospital 1 |  |  | Hospital 2 |  |  | Both hospitals |  |  |
| --- | --- | --- | --- | --- | --- | --- | --- | --- | --- |
|  | Total | Seropositive |  | Total | Seropositive |  | Total | Seropositive |  |
|  | N | n | % | N | n | % | N | n | % |
| Domestic/ Cleaning | 62 | 15 | 24% | 45 | 9 | 20% | 107 | 24 | 22% |
| Catering | 83 | 23 | 28% | 18 | 5 | 28% | 101 | 28 | 28% |
| Maintenance | 27 | 6 | 22% | 24 | 1 | 4.2% | 51 | 7 | 14% |
| Security | 33 | 6 | 18% | 14 | 3 | 21% | 47 | 9 | 19% |
| Porter | 20 | 6 | 30% | 13 | 1 | 7.7% | 33 | 7 | 21% |
| Chaplain | 11 | 3 | 27% | 2 | 0 | 0.0% | 13 | 3 | 23% |
| Other | 7 | 1 | 14% | 0 | 0 | - | 7 | 1 | 14% |
| Driver | 0 | 0 | - | 6 | 2 | 33% | 6 | 2 | 33% |
| Total | 243 | 60 | 25% | 122 | 21 | 17% | 365 | 81 | 22% |

**Table 2h.** Prevalence of asymptomatic SARS-CoV-2 infection, by hospital location and ethnicity, April 2021

|  |  | Total<br>seropositive | Asymptomatic SARS-CoV-2 infection |  |  |
| --- | --- | --- | --- | --- | --- |
|  |  | N | n | % (95% CI) | p-value* |
| **St James's Hospital** | Total | 623 | 121 | 19 (16 - 23) | 0.009 |
|  | Irish | 401 | 82 | 20 (17 - 25) |  |
|  | Other white background | 56 | 13 | 23 (13 - 36) |  |
|  | Asian | 122 | 13 | 11 (5.8 - 18) |  |
|  | African or other black background | 29 | 11 | 38 (21 - 58) |  |
|  | Other | 15 | 2 | 13 (1.7 - 40) |  |
| **Galway University Hospital** | Total | 275 | 48 | 17 (13 - 22) | 0.531 |
|  | Irish | 194 | 35 | 18 (13 - 24) |  |
|  | Other white background | 38 | 5 | 13 (4.4 - 28) |  |
|  | Asian | 26 | 5 | 19 (6.6 - 39) |  |
|  | African or other black background | 10 | 3 | 30 (6.7 - 65) |  |
|  | Other | 7 | 0 | - |  |
| **Both hospitals** | Total | 898 | 169 | 19 (16 - 22) | 0.010 |
|  | Irish | 595 | 117 | 20 (17 - 23) |  |
|  | Other white background | 94 | 18 | 19 (12 - 29) |  |
|  | Asian | 148 | 18 | 12 (7.4 - 19) |  |
|  | African or other black background | 39 | 14 | 36 (21 - 53) |  |
|  | Other | 22 | 2 | 9.1 (1.1 - 29) |  |

\*Calculated using the Chi-square test

**Table 2i.** Undiagnosed SARS-CoV-2 infection, by HCW role and hospital location, April 2021

|  | St James's Hospital |  | University Hospital Galway |  | Both hospitals |  |
| --- | --- | --- | --- | --- | --- | --- |
|  | n | % | n | % | n | % |
| **Admin** | 19 | 10% | 5 | 11% | 24 | 10% |
| **Medical/dental** | 12 | 6.3% | 8 | 18% | 20 | 8.5% |
| **Nursing/ midwifery** | 79 | 42% | 19 | 42% | 98 | 42% |
| **Allied health** | 32 | 17% | 6 | 13% | 38 | 16% |
| **General support** | 22 | 12% | 4 | 8.8% | 26 | 11% |
| **Health care assistant** | 26 | 14% | 2 | 4.4% | 28 | 12% |
| **Other** | 0 | 0.0% | 1 | 2.2% | 1 | 0.4% |
| **Total** | 190 | 100% | 45 | 100% | 235 | 100% |

**Table 3b.**
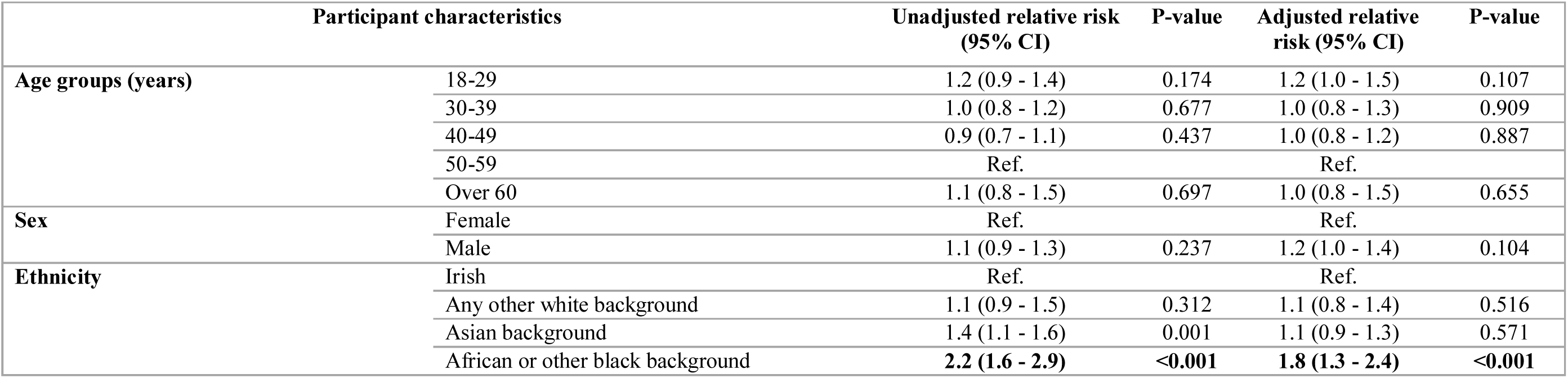
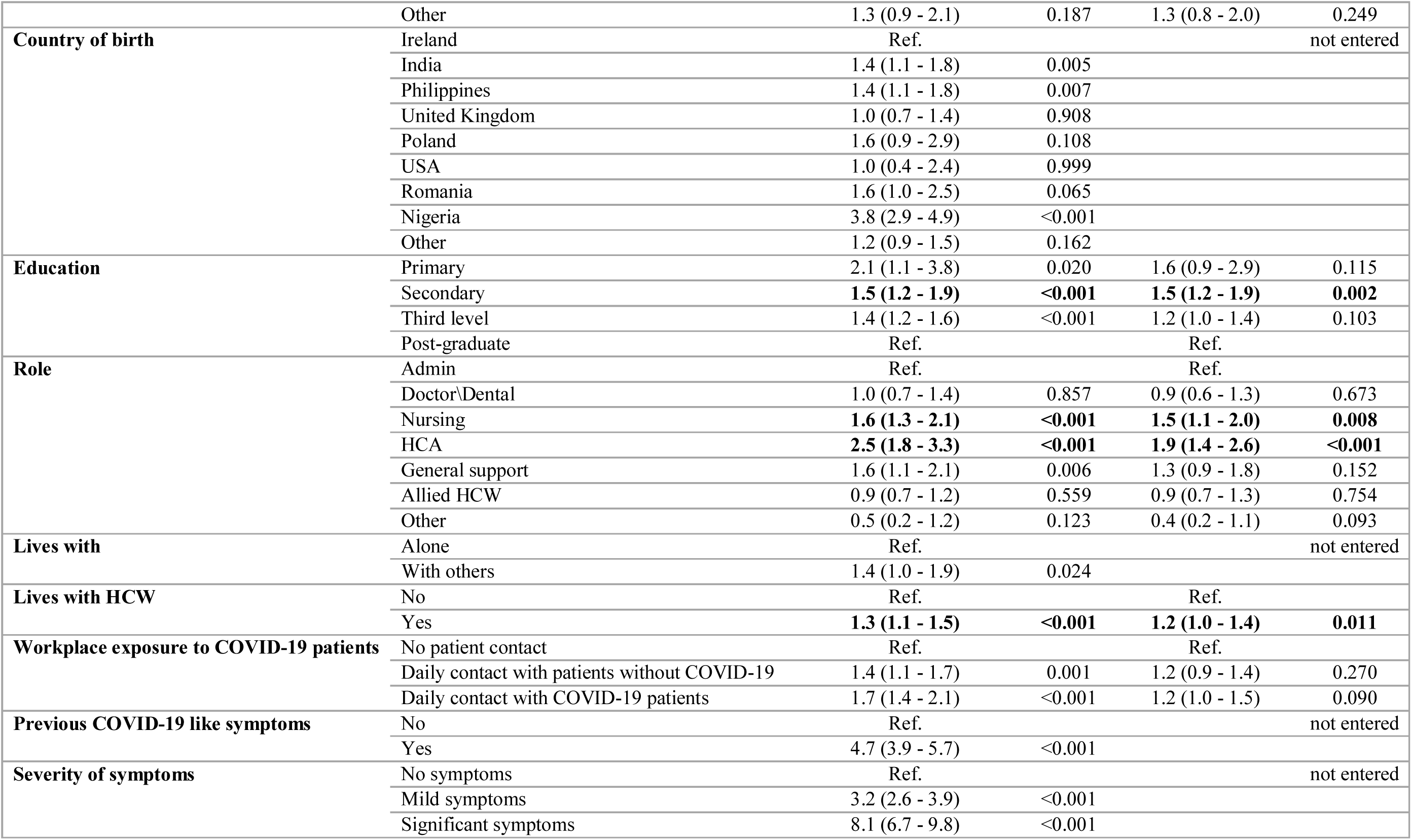
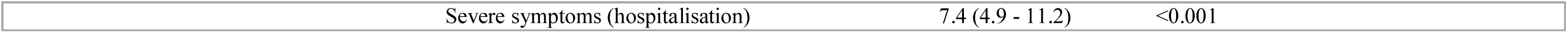
Association between risk factors and SARS-CoV-2 seropositivity, Hospital 1, April 2021

**Table 3c.**
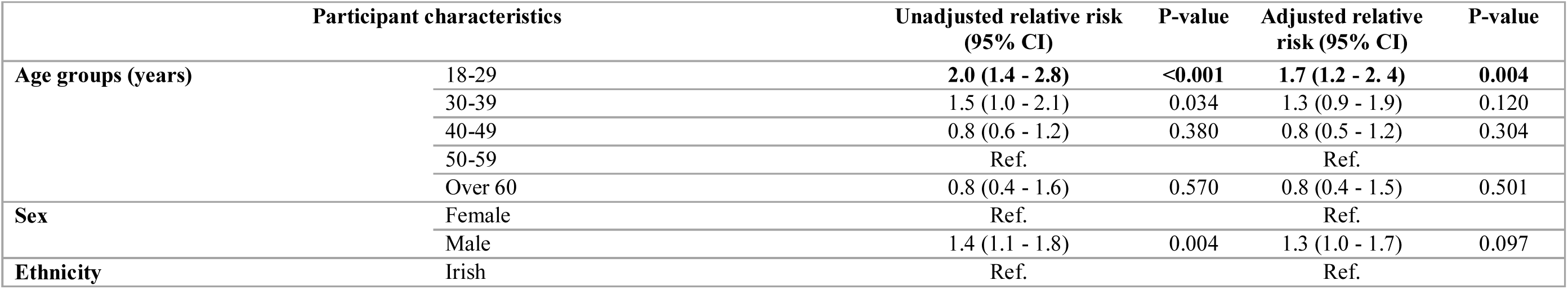
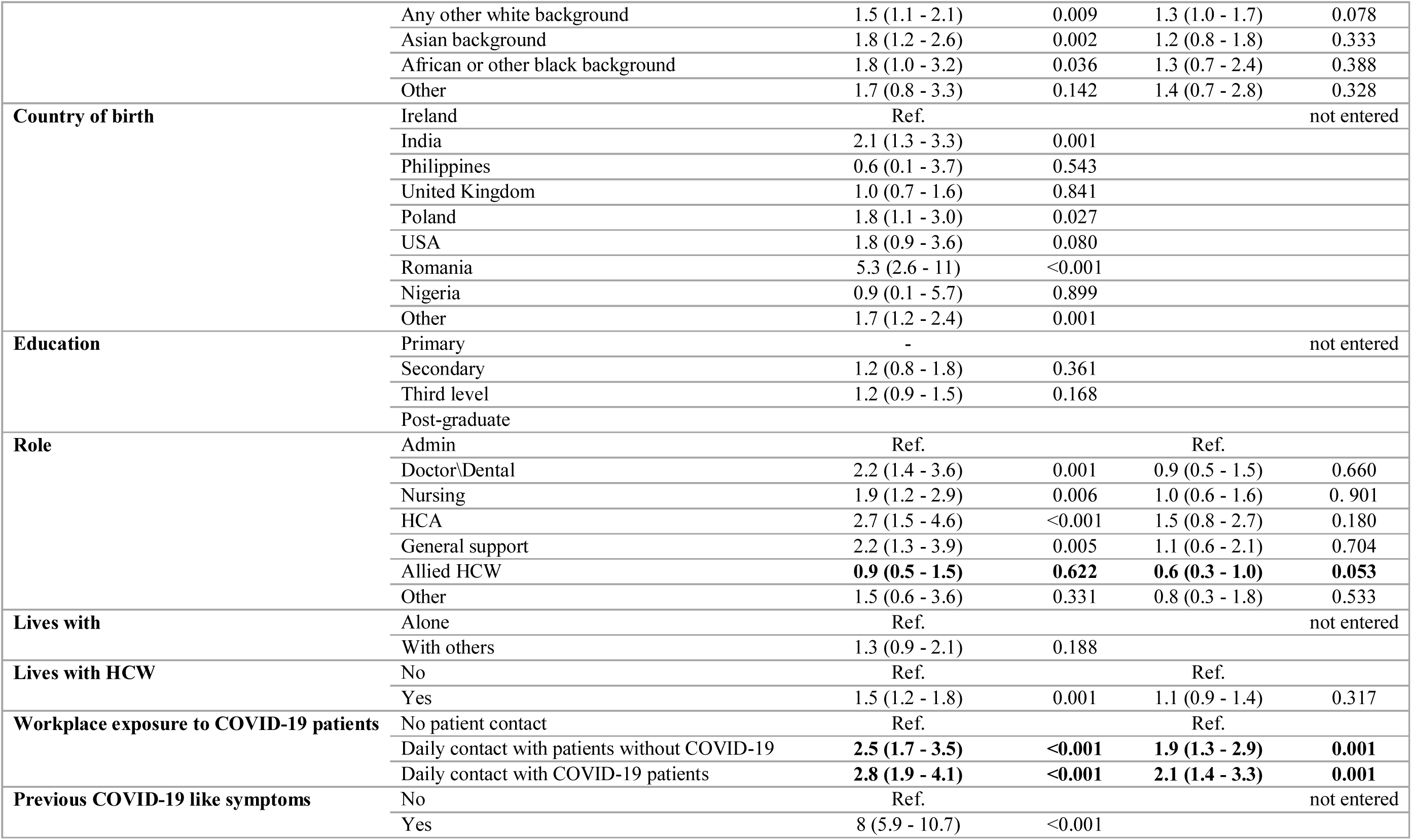
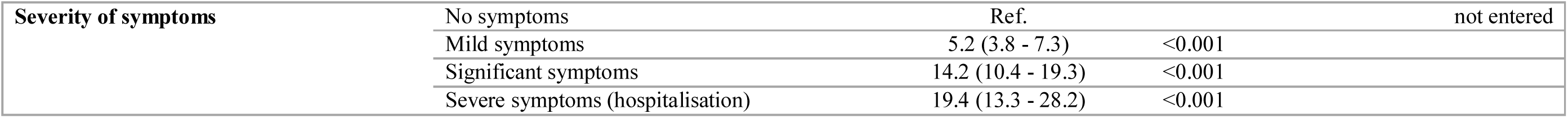
Association between risk factors and SARS-CoV-2 seropositivity, Hospital 2, April 2021

**Table F.**
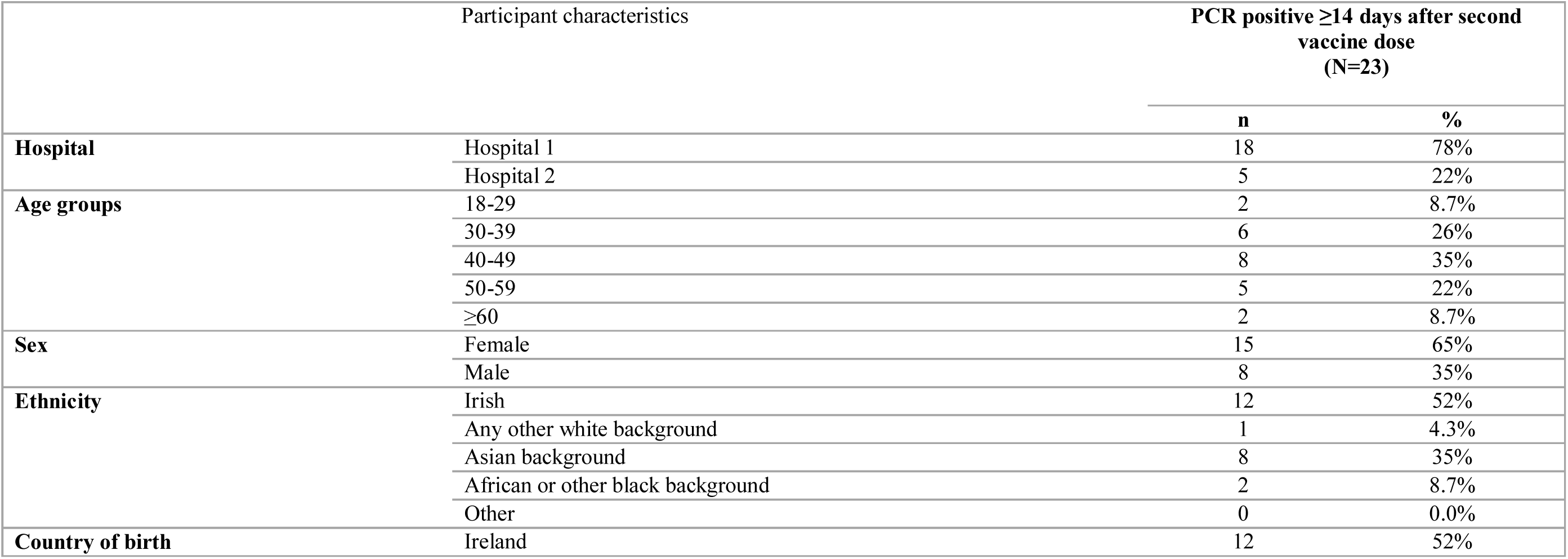
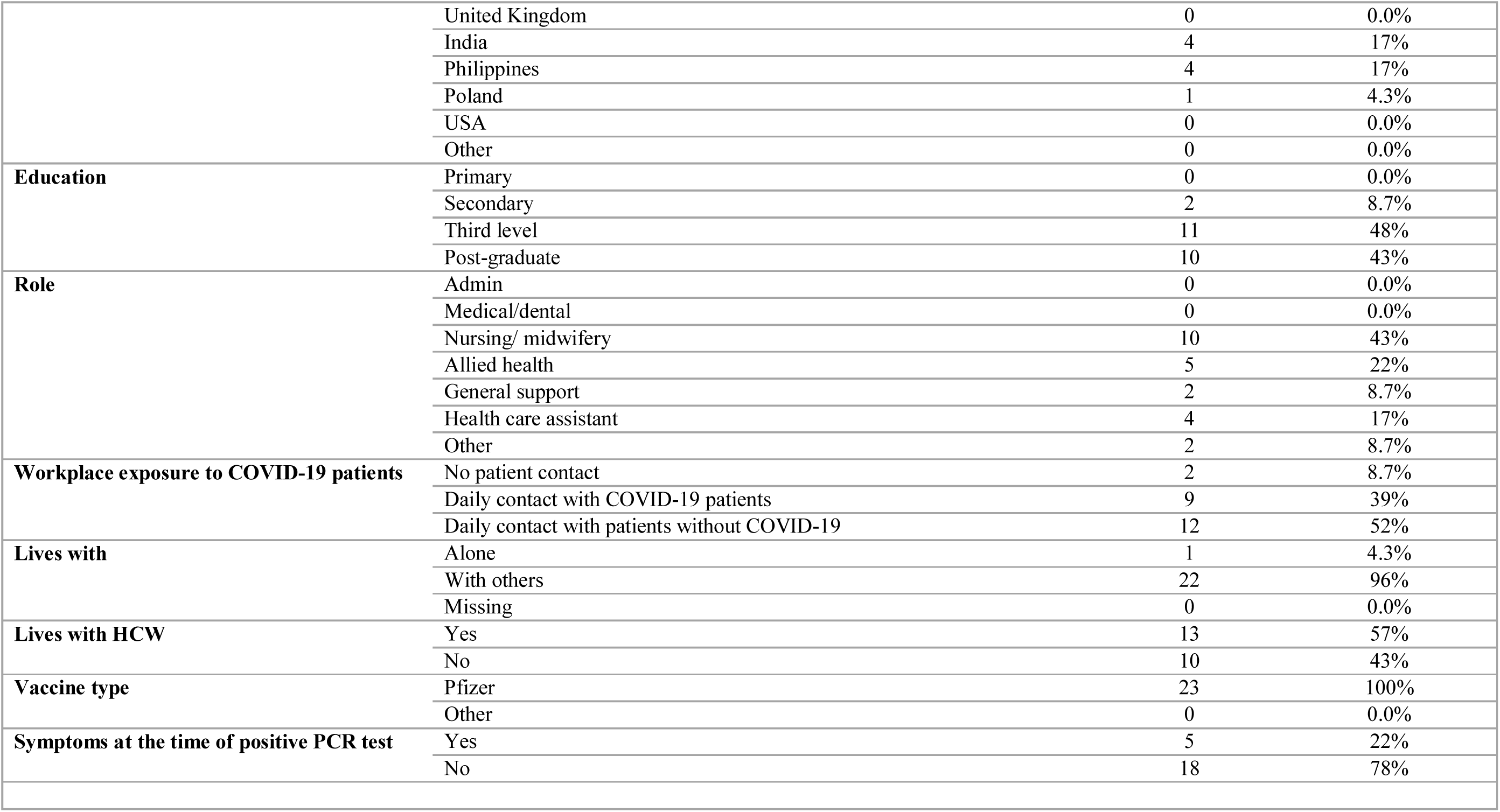
Characteristics of fully vaccinated participants with PCR- confirmed infection i.e. vaccine breakthrough cases, both hospitals (n=23), PRECISE 2, April 2021

| Participant characteristics |  | PCR positive ≥14 days after second vaccine dose (N=23) |  |
| --- | --- | --- | --- |
|  |  | n | % |
| **Hospital** | Hospital 1 | 18 | 78% |
|  | Hospital 2 | 5 | 22% |
| **Age groups** | 18-29 | 2 | 8.7% |
|  | 30-39 | 6 | 26% |
|  | 40-49 | 8 | 35% |
|  | 50-59 | 5 | 22% |
|  | ≥60 | 2 | 8.7% |
| **Sex** | Female | 15 | 65% |
|  | Male | 8 | 35% |
| **Ethnicity** | Irish | 12 | 52% |
|  | Any other white background | 1 | 4.3% |
|  | Asian background | 8 | 35% |
|  | African or other black background | 2 | 8.7% |
|  | Other | 0 | 0.0% |
| **Country of birth** | Ireland | 12 | 52% |
|  | United Kingdom | 0 | 0.0% |
|  | India | 4 | 17% |
|  | Philippines | 4 | 17% |
|  | Poland | 1 | 4.3% |
|  | USA | 0 | 0.0% |
|  | Other | 0 | 0.0% |
| **Education** | Primary | 0 | 0.0% |
|  | Secondary | 2 | 8.7% |
|  | Third level | 11 | 48% |
|  | Post-graduate | 10 | 43% |
| **Role** | Admin | 0 | 0.0% |
|  | Medical/dental | 0 | 0.0% |
|  | Nursing/ midwifery | 10 | 43% |
|  | Allied health | 5 | 22% |
|  | General support | 2 | 8.7% |
|  | Health care assistant | 4 | 17% |
|  | Other | 2 | 8.7% |
| **Workplace exposure to COVID-19 patients** | No patient contact | 2 | 8.7% |
|  | Daily contact with COVID-19 patients | 9 | 39% |
|  | Daily contact with patients without COVID-19 | 12 | 52% |
| **Lives with** | Alone | 1 | 4.3% |
|  | With others | 22 | 96% |
|  | Missing | 0 | 0.0% |
| **Lives with HCW** | Yes | 13 | 57% |
|  | No | 10 | 43% |
| **Vaccine type** | Pfizer | 23 | 100% |
|  | Other | 0 | 0.0% |
| **Symptoms at the time of positive PCR test** | Yes | 5 | 22% |
|  | No | 18 | 78% |

**Table 4.**
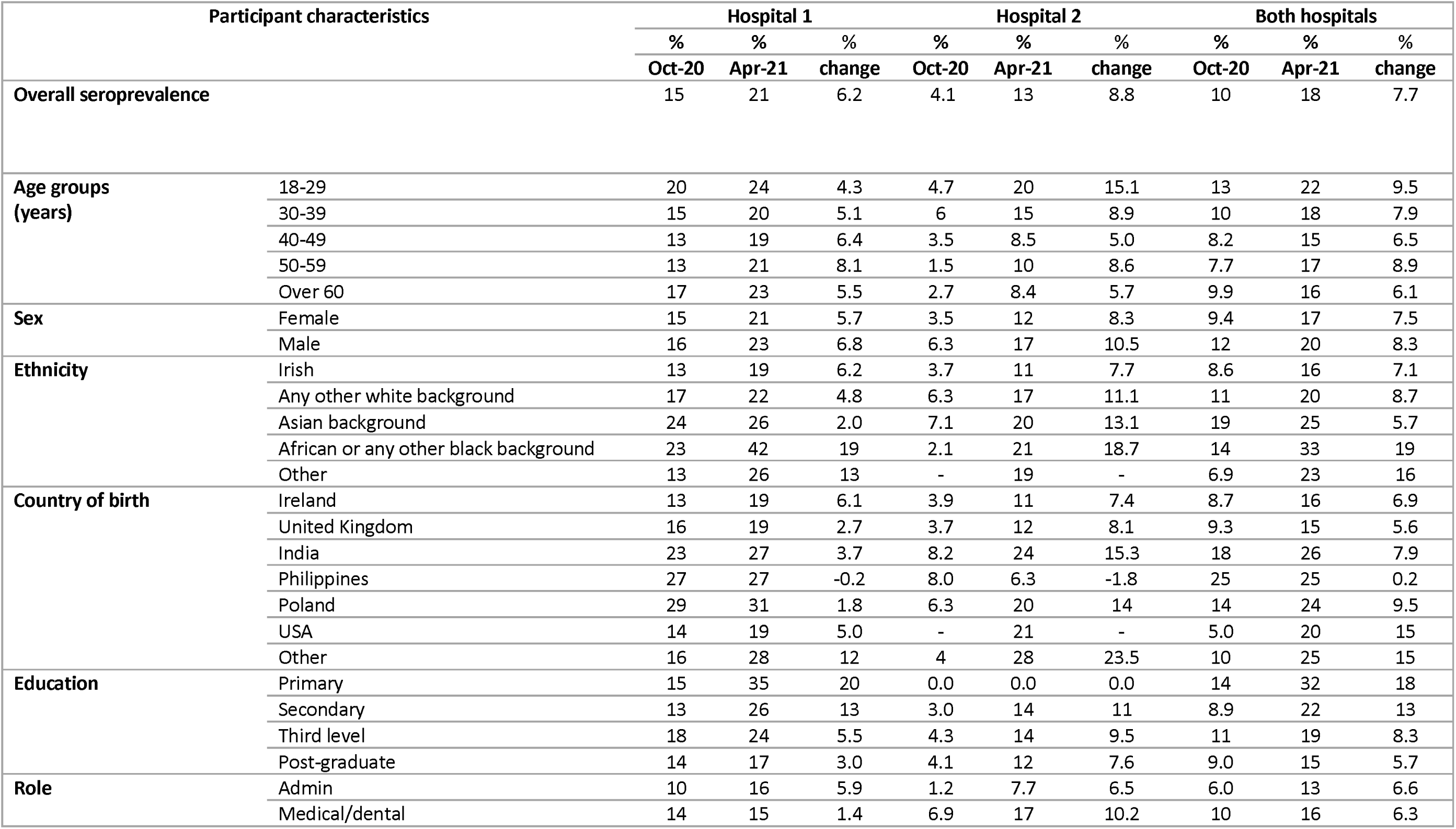
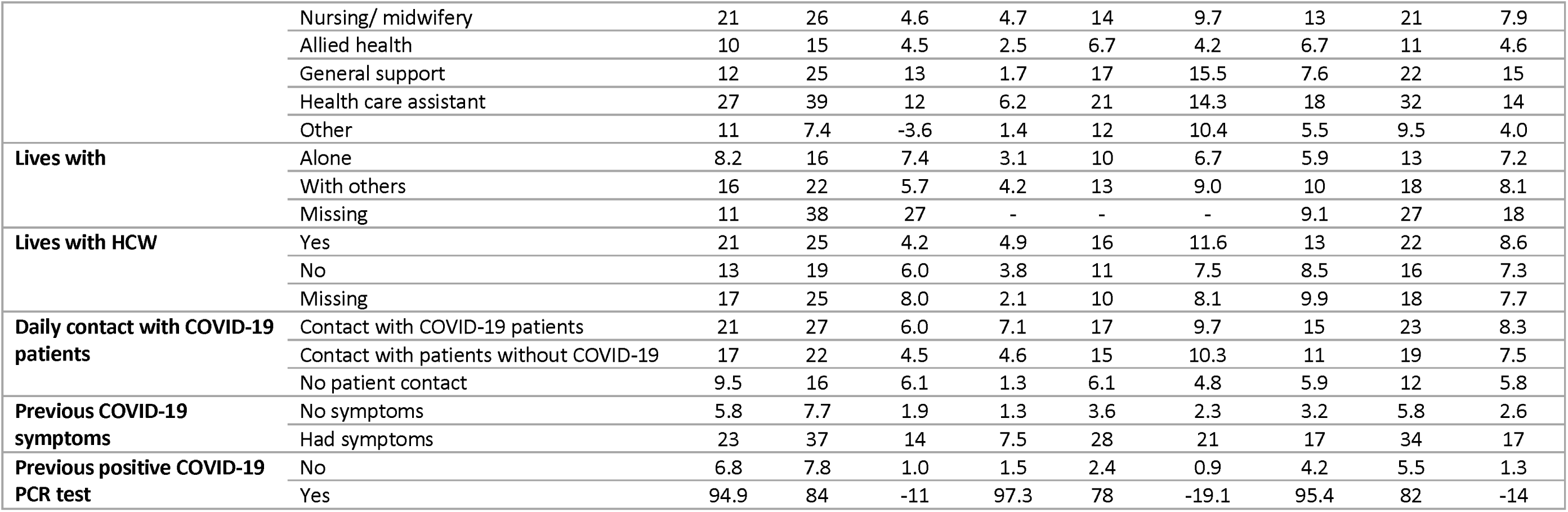
Comparison of SARS-CoV-2 seroprevalence October 2020 and April 2021, by hospital

